# Knowledge, Attitude, and Practice (KAP) towards Rabies, its Suspected Cases and Associated Economic Impact in West Shewa Zone, Oromia, Ethiopia

**DOI:** 10.1101/2024.10.24.24316052

**Authors:** Tegegn Dilbato, Nugusa Desalegn, Tolesa Negasa, Gudina Mekonnin, Kebede Abdisa, Hana Dejene, Segni Bedasa, Moges Kidane, Endrias Zewdu

## Abstract

**Background:** Rabies is a neglected zoonotic disease that affects many developing countries in which it is endemic, including Ethiopia. Regardless of its endemic/epidemic patterns, no study has been conducted on rabies in the study area.

**Methods:** A cross-sectional, and retrospective study designs were carried out in selected districts of the West Shewa zone to assess the knowledge, attitude, and practice and estimate the economic impacts of rabies in humans and animals from 2017 to 2021. A total of 422 informants were selected as study participants through simple random sampling techniques from the community. A pre-tested semi-structured questionnaire was used to gather information from the participants. The relevant data was analyzed using SPSS version 20 and then described and interpreted using descriptive statistics and a binary logistic regression test.

**Results:** The results indicated that the community had 91.2%, 74.2%, and 81% knowledge, attitude, and practice scores on rabies. Having a dog (AOR=1.7, 95% CI: 1.050, 2.87), acquiring information from mass media (AOR=0.4, 95% CI: 0.175, 0.788; P-value <0.05), and having a history of previous exposure (AOR=7.3, 95% CI: 1.618, 32.69) were significantly associated with knowledge score about rabies. Positive attitudes toward rabies were higher in dog owners (AOR=2.5, 95% CI: 1.074, 5.993) and private workers (AOR=9.5, 95% CI: 1.981, 45.76). Living in an urban area (AOR=0.19, 95% CI: 0.044, 0.881) and Dirre Inchinni district (AOR=0.03, 95% CI: 0.07, 0.150) had good practices level of rabies. In addition, 579 suspected human rabies cases and 183 animal cases were registered during the study periods. The number of rabies cases that occurred in humans, was higher than in animals in the study area, with varying degrees of incidence. Rabies in the study area resulted in direct and indirect costs of around 142590 USD.

**Conclusions:** The study showed that most of the respondents were unaware of rabies. Therefore, effective and well-organized control measures like annual vaccination and awareness campaigns should be developed, focusing on identified risk factors and areas. The incidence of the diseases should be considered while designing control and prevention measures.

**Author Summary:** Rabies is a life-threatening zoonotic disease that poses a serious public health and socio-economic impact, especially in regions with poor access to healthcare services and vaccination. The study aimed to assess the community’s awareness of rabies management and prevention and determine the number of suspected cases and the associated economic impact. It assessed what the community knew about the symptoms, prevention, and transmission of rabies from animals to humans. It also identified attitudes that were commonly embraced regarding vaccination and post-exposure prophylaxis (PEP), how people handled dog bites, and the best courses of action. The suspected cases and the economic impact of rabies were determined from registered data on human and animal cases in medical casebooks and veterinary clinics. Because of financial and logistical difficulties like tight budgets and shortages of supplies in the region, the victim households travel a long distance to hospitals to get post-exposure prophylaxis (PEP). Both direct costs (such as medical care and post-exposure prophylaxis) and indirect costs (such as costs for public health, animal management, travel costs, lost wages, and other alternative medications for adverse reactions) were considered to estimate the economic impact. The economic burden on humans was approximated due to the absence of reliable animal rabies data. This study also offered a systematic analysis of how rabies impacts the livelihoods of West Shewa’s rural and urban communities. Furthermore, the study also underlined the significance of responsible dog ownership, the substantial economic impact of rabies in the region, and crucial insights into the gaps in knowledge and practices related to rabies control. By giving policymakers, medical experts, and veterinarians vital information to establish dedicated actions addressing One health approach to reduce rabies transmission and its socioeconomic effects, our study supports the larger regional effort to combat the disease. Enhancing rabies control and prevention strategies, like raising vaccination coverages, and awareness campaigns, strengthening community involvement, and establishing robust rabies surveillance systems in control programs should be recommended.

## Introduction

Rabies is a zoonotic viral disease caused by a virus belonging to the Lyssavirus genus of the Rhabdoviridae family and Mononegavirales order [1]. This virus infects mammals and causes lethal encephalitis [2]. Rabies affects all warm-blooded animals, including humans, and is common in many locations, including Ethiopia. It is distributed worldwide, and the most affected countries are tropical Africa and Asia. The disease is of concern mainly because of its zoonotic importance [3].

It is a life-threatening zoonotic disease transmitted through carnivores to humans and livestock through biting. It is known to cause significant morbidity and mortality among humans and animals each year. Almost half of cases occur in children under the age of 15 years [4,5]. In humans, the incubation period of rabies is highly variable, ranging from days to years, with an average of 2–3 months, being influenced by: (i) location, extent, and depth of the wound; (ii) distance between the location of the wound and the central nervous system; (iii) concentration of inoculated virus particles, and (iv) virus strain. The incubation period in children tends to be of shorter duration than adults [1,6].

Rabies is a tropical neglected, highly prioritized disease because it affects developing countries. Despite it causing a massive loss in the livestock industry in Ethiopia, only less attention is given to it [7]. The knowledge, attitude, and practice survey done in the country shows there is still a gap in disease control and prevention [8]. Rabies is considered a non-transmitted disease with little public awareness; even though these diseases are among the oldest, the challenges associated with their prevention in both animals and people remain unresolved. The contribution of human attitudes and behavior to prevention is poorly understood [9]. Ethiopia is one of the worst affected countries with rabies, in which domestic dogs are the primary source of human infection [10].

In Ethiopia, few studies have been conducted to assess community awareness towards rabies, concentrated in Addis Ababa due to the large population of stray dogs. Poor public awareness of rabies is considered one of the bottlenecks for the prevention and control of rabies in Ethiopia, especially in rural areas [8].

The reliability of the reported incidence data differs among Ethiopia’s regions due to geographical and cultural differences. For instance, in rural Ethiopia, individuals exposed to rabies often prefer to see traditional healers to diagnose and treat the disease because of cultural background, lack of knowledge, or limited accessibility to medical treatment. These widespread traditional practices of handling rabies cases might interfere with medical-treatment-seeking practices due to strong belief in traditional medicines, resulting in an underreporting of the actual number of rabies cases and their related health burden. Even though rabies in wild and domestic animals poses a threat to human life and significant damage due to livestock loss, few studies to date examine the economic impact of rabies on humans and animals [11,12,13].

Due to the lack of appropriate registration systems, accurate data on the incidences of rabies in humans, dogs, and livestock are rare in Ethiopia. Medical and veterinary records on rabies are insufficient and cannot be utilized to accurately measure the disease’s impact [14,15]. Even if there is a high prevalence of rabies throughout the world, including Ethiopia, there is still a gap in knowledge, attitudes, practices, and economic impacts of rabies on animals and humans. In addition, there is no empirical studies on direct or indirect economic costs associated with human exposure to rabies have been conducted in the study area. The results of an interview and archive study conducted in two districts to estimate the direct and indirect costs of human exposure to probable rabies are presented in this article. There is a scarcity of information on rabies prevention and control practices and associated factors among dog owners in Ethiopia on rabies. Knowledge of responsible dog ownership and dog population management among the public is low. Therefore, the study aims to assess knowledge, attitude, and practice (KAP), to determine the incidence and to evaluate economic effects of rabies on humans in the selected districts of West Shoa, Ethiopia.

## Methods

### Ethical consideration

The study’s scientific soundness and rationale were approved by the Ethical and Research Review Board of Ambo University. Before data collection, a brief discussion was conducted with respondents about the objective of the study and obtained their verbal consent for collaboration. There are no animal subjects included in this study.

### Study location

This study was conducted in two selected districts of West Shewa Zone, Oromia Regional State, Ethiopia, namely: the Dire Inchini and Ambo districts. West Shewa Zone is located between Latitude 9° 09’ 60.00” N and Longitude 37° 49’ 59.99” E with a maximum elevation of 3,337m and an average elevation of 1,811m. West Shewa is bordered on the south by the Southwest Shewa Zone and the Southern Nations, Nationalities, and Peoples Region, on the southwest by the Jimma Zone, on the west by the East Welega Zone, on the northwest by Horo Gudru Welega Zone, on the north by the Amhara Region, on the northeast by North Shewa, and the East by Oromia Special zone Surrounding Finfinnee. Its highest point is Mount Wenchi (3386 meters); other notable peaks include Mount Mengesha and Mount Wechacha. The zone has a total population of 2,058,676, of whom 1,028,501 are men and 1,030,175 women. A total of 428,689 households were counted in this Zone, which results in an average of 4.80 persons per household and 415,013 housing units. The animal population in the West Shewa zone is as follows: 3,795,500 cattle, 1,262,284 sheep, 829,046 goats, 271,391 horses, 250,155 donkeys, 21,836 mules, and 209,5500 chicken. The map of the study area districts is indicated in (**Fig. 1**).

**Fig. 1:**
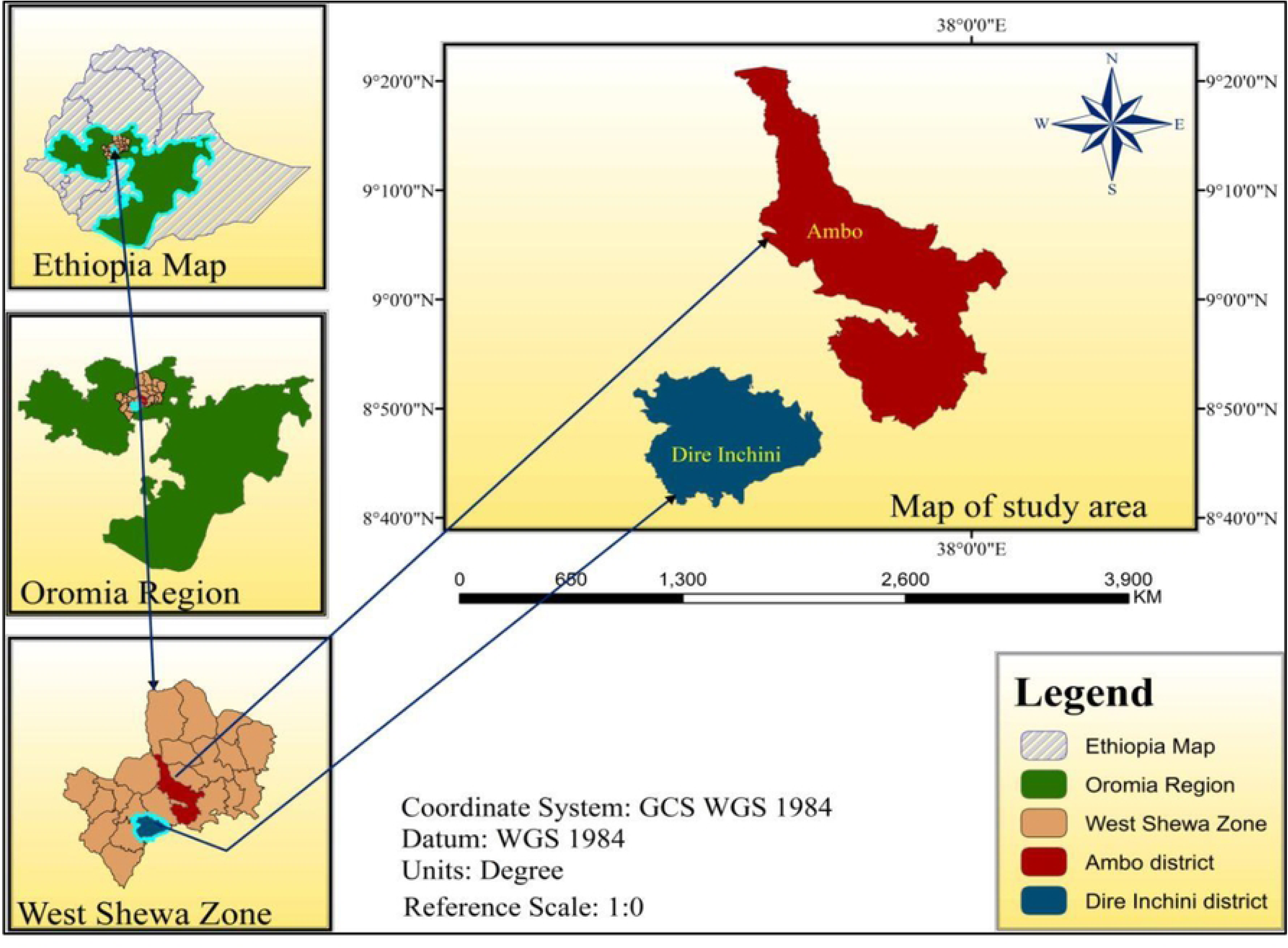
Map of the study area Source: (Arc GIS, Version 14)

### Study population

The study populations were humans and animals found in two selected districts of the study, including adult community members aged 18 years and above, veterinary workers, and health workers who had lived in districts for more than one year to obtain complete information before the commencement of this survey. A total of 422 respondents were included in the study to assess their knowledge, attitude, and practice. The study used all the data about rabies obtained from those health centers to determine economic burdens.

### Study designs

Both cross-sectional and retrospective study designs were used to assess the knowledge, attitudes, practice, and economic burdens of rabies in animals and humans among community members in selected districts of the West Shewa zone. The collected data was used to compare the community’s knowledge, attitude, and practice and the economic effects of rabies in animals and humans. About six key informants from the community were included in the survey while collecting the information with a questionnaire. Those respondents were veterinary workers, health workers, local leaders, an older man, and community farmers.

### Sampling method and sample size calculation

In this study, the districts were selected using a purposive sampling technique based on the burden of the disease. In contrast, the respondents were selected from the population through the random sampling method. The required sample size for this study was estimated by considering the proportion score of knowledge 46.1%, attitude 56.5%, and practice 63.5% [16] with a 5% margin of error and 95% confidence level. Thus, the calculated sample sizes for knowledge, attitude, and practice were 382, 384, and 356, respectively. The larger sample size among the knowledge, attitude, and practice is taken as appropriate, which is 384. Therefore, the total sample size was computed by adding a 10% non-response rate; thus, the total sample size was 422 respondents. The study respondents were selected from the existing households (2573 households) in the selected peasant associations (PAs) depending on their total households proportionally (**Fig.2**).

**Fig 2.**
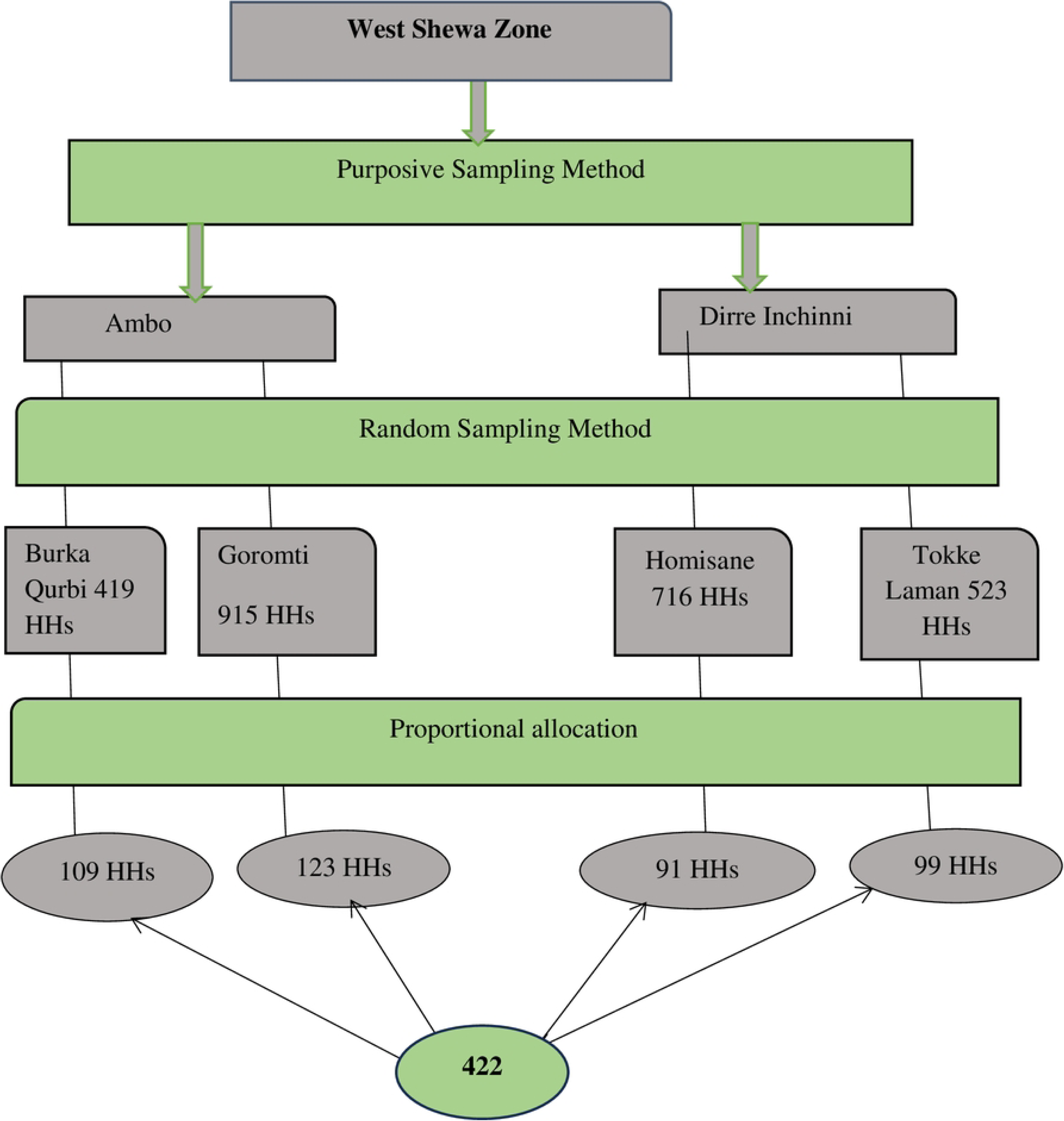
A conceptual framework for informants’ selection

**Fig 3.**
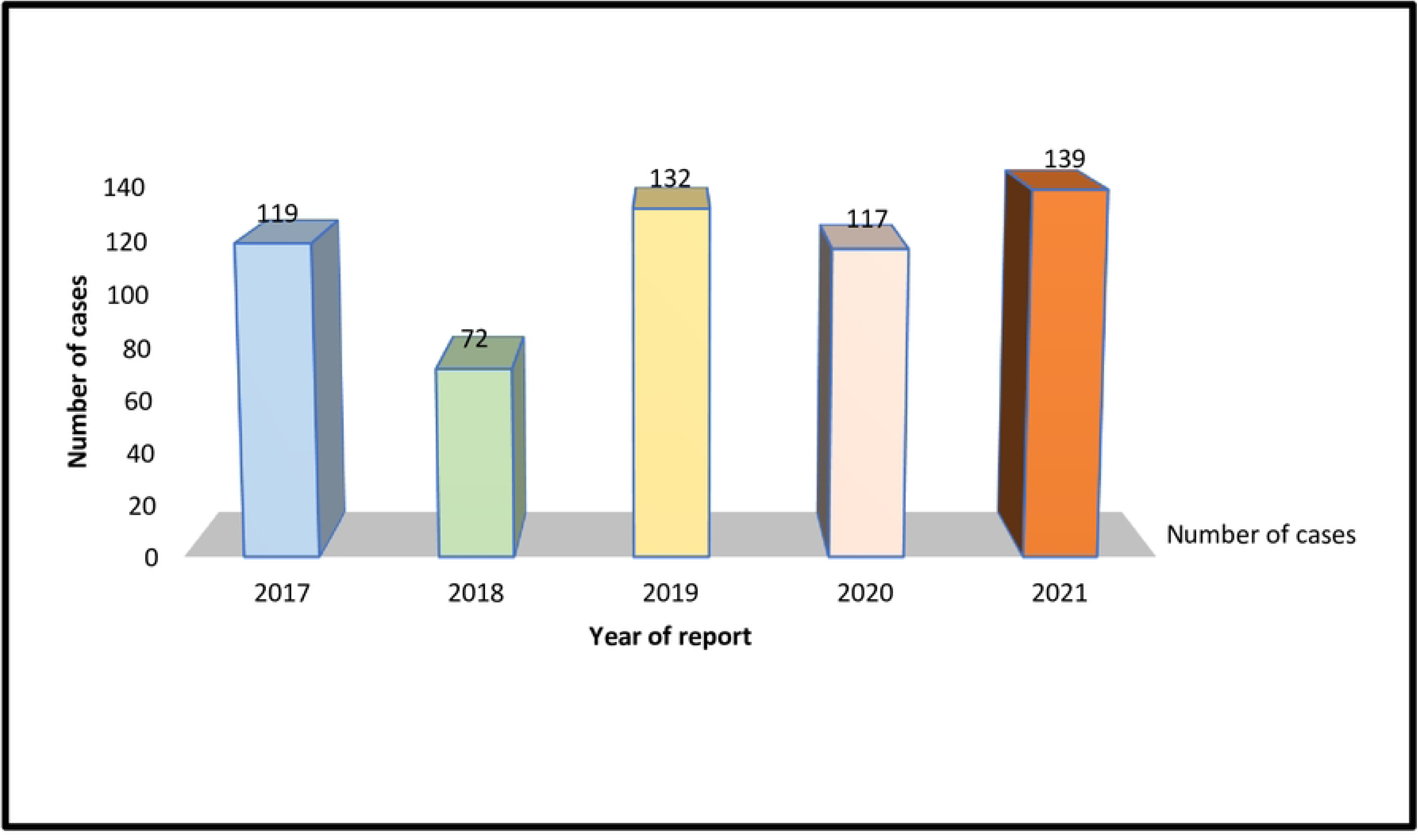
Temporal distribution of rabies in the study area

## Methods of data collection

### Retrospective data

Retrospective data were collected from individual patients taking rabies post-exposure prophylaxis at the health centers and veterinary clinics during the study period. The data collected was used to compare the level of the cases and the economic impacts of rabies in the community.

### Questionnaire survey data

Semi-structured questionnaires were used for data collection from community respondents of the survey areas between 2017 and 2022. The questionnaire was prepared in English and later translated into the local language (Afaan Oromo). The relevant data captured included socio-demographic variables, knowledge, attitudes, practices, and economic impacts of rabies on animals and humans. It consists of five segments: (1) items regarding the respondent’s socio-demographic information (address, age, sex, education level, occupation, animal ownership status); (2) questions related to the knowledge of rabies; and (3) questions related to attitudes of rabies; and (4) questions related to the practice of rabies and finally item consists (5) questions related to economic impacts of rabies on humans. The questionnaire was previously assessed to improve clarity and interpretation. For the knowledge assessment, 11 questions, attitude, 7 questions, and practice 9 questions were used for the KAP assessment for rabies.

### Data management and analysis

The primary data collected from households through questionnaires was entered into a Microsoft Excel spreadsheet, checked, filtered, scored, and further exported to SPSS version 20 for descriptive analysis to summarize the data (percentage and frequency distribution). The data complimented, elaborated quantitative findings, and clarified relevant aspects of rabies-related knowledge, attitudes, practices, and economic impacts. Logistic regression was utilized to build a model to identify and quantify associations. P-value was used to assess if there was an association between risk factors (age, sex and education status, religion, source of information, previous exposure, marital status, and occupation) which are independent variables, and KAP scores (dependent variable). In all cases, a 95% CI was employed to estimate sample results for the target population in the study area. Values of P<0.05 were considered statistically significant. Knowledge, attitude, and practice scores were classified as “poor” for those who scored below 50% and “good” for those who scored above 50%.

The total cost (TC) per suspected human exposure to rabies was calculated by adding the direct costs (DC) and indirect costs (IC) of rabies-related charges paid by patients, district animal control units, and public health services. The total cost of rabies in humans was estimated using the method described by [17,18] considering the different costs paid by patients. The following formula, where *pep* is the cost of PEP and *med* is the cost of other medical expenses, was used to estimate TC and IC.

Direct cost (DC) = *pep* + *med*

Where, *pep* is the cost of PEP, and *med* is other medical costs.

Indirect cost (IC) = *ph* + *ac* + *o* + *tt* + *iw* + *alt*

Where *ph* is entailed public health costs, *ac* is animal control costs, *o* is other indirect costs, tt is travel time costs, *lw* is lost wages, and *alt* is alternative medicines for adverse reactions.

Total cost (TC) = *DC*+*IC* = pep + med + ph + ac + o + tt + iw + alt

## Results

### Demographic characteristics of study participants

The socio-demographic profiles of the respondents are presented in (**Table 1**). Out of 422 participants, most of the respondents who participated in the survey were from the Ambo district, 232 (55%), and the remaining 190 (45%) were from the Dirre Inchinni district, respectively. Of the total participants, 376 (89.1%) were males, while 46 (10.9%) were females, and most of them, 274 (49.9 %), lived in rural areas.

**Table 1.** Socio-demographic profiles of the respondents in the study area.

| **Variables** | **Category** | **No of respondents** | **Percentage (%)** |
| --- | --- | --- | --- |
| **Dog ownership** | No | 332 | 78.7 |
|  | Yes | 90 | 21.3 |
| **District** | Ambo | 232 | 55.0 |
|  | Dirre Inchinni | 190 | 45.0 |
| **Residence** | Peri-urban | 60 | 14.2 |
|  | Urban | 88 | 20.9 |
|  | Rural | 274 | 64.9 |
| **Age** | 18-30 | 49 | 11.6 |
|  | 31-50 | 272 | 64.5 |
|  | 51-70 | 101 | 23.9 |
| **Sex** | Female | 46 | 10.9 |
|  | Male | 376 | 89.1 |
| **Marital status** | Divorced | 2 | 0.5 |
|  | Widowed | 2 | 0.5 |
|  | Single | 4 | 4.0 |
|  | Married | 401 | 95.0 |
| **Level of education** | Illiterate | 203 | 48.1 |
|  | Primary | 113 | 26.8 |
|  | Secondary | 65 | 15.4 |
|  | Above grade12 | 41 | 9.7 |
| **Religion** | Muslim | 33 | 7.8 |
|  | Orthodox | 204 | 48.3 |
|  | Protestant | 185 | 43.8 |
| **Occupation** | Gov't Employee | 20 | 4.7 |
|  | Private worker | 10 | 2.4 |
|  | Farmer | 392 | 92.9 |
| **Source of information** | Family/Friends | 338 | 80.1 |
|  | Mass media | 23 | 5.5 |
|  | Mixed source | 51 | 12.1 |
| **Previous exposure** | Yes | 297 | 70.4 |
|  | No | 125 | 29.6 |

### Knowledge, attitude, and practice score and risk factors on rabies

Of all study participants, most of the respondents, 395 (93.6%), have ever heard of the disease called rabies, which is locally called ‘’Dhukkuba Saree Maraatuu’’. When asked to mention the animals commonly affected by rabies, 302 (71.6%) of the respondents reported dogs, followed by cattle 79 (18.7%), as indicated in (**Table 2)**.

**Table 2.**
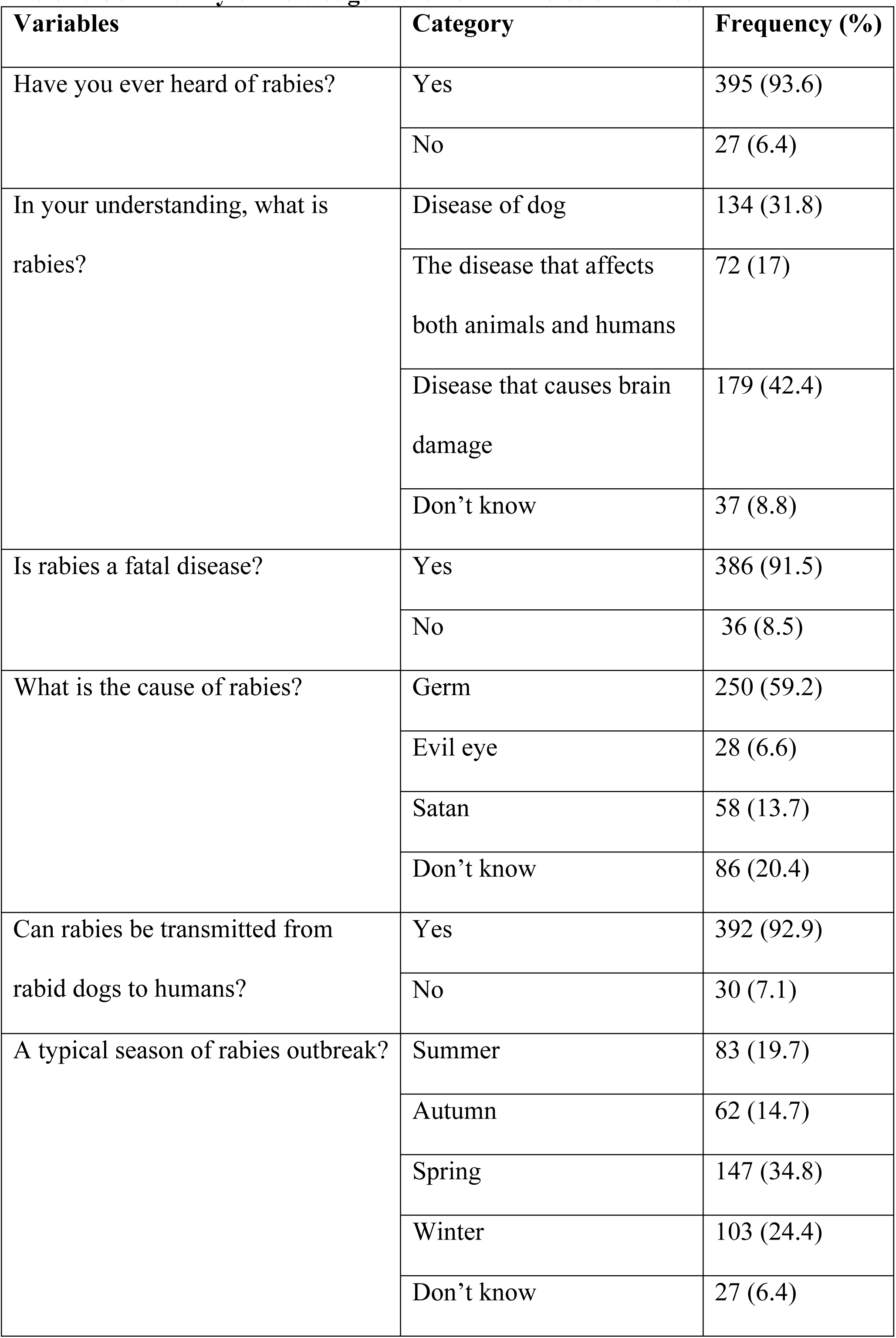
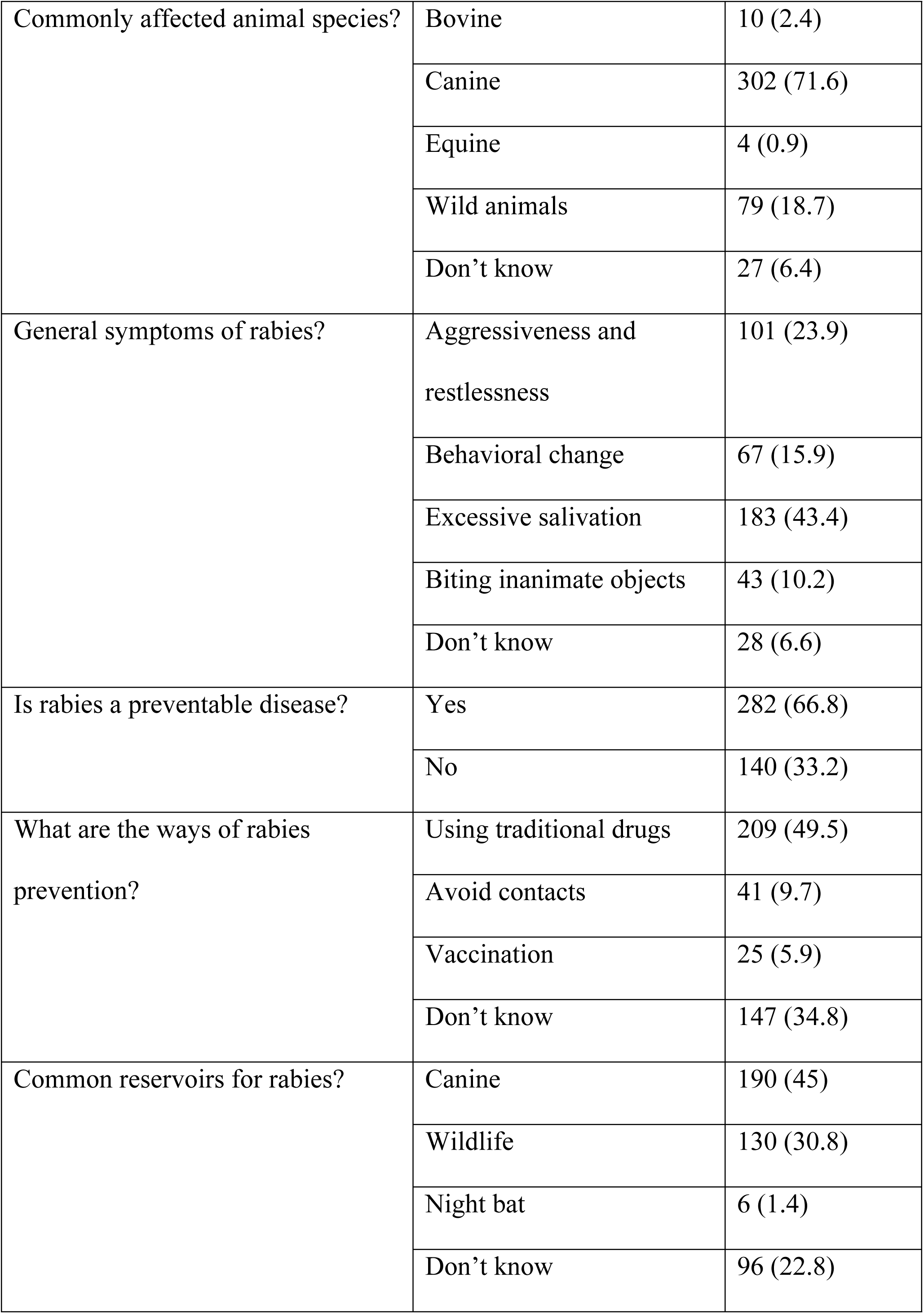
Community’s knowledge indicator variables on rabies.

Table (3) depicts the logistic regression analysis results between knowledge score and risk factors on rabies. The study revealed that more than half of the respondents, 385 (91.2%), had good knowledge about rabies. The association between independent variables and knowledge score on rabies was analyzed using logistic regression. There was a significant association between knowledge score and dog ownership (AOR=1.7, 95% CI: 1.050, 2.873), had previous exposure to rabies (AOR=7.3, 95 CI: 1.62, 32.69) with knowledge score.

**Table 3.**
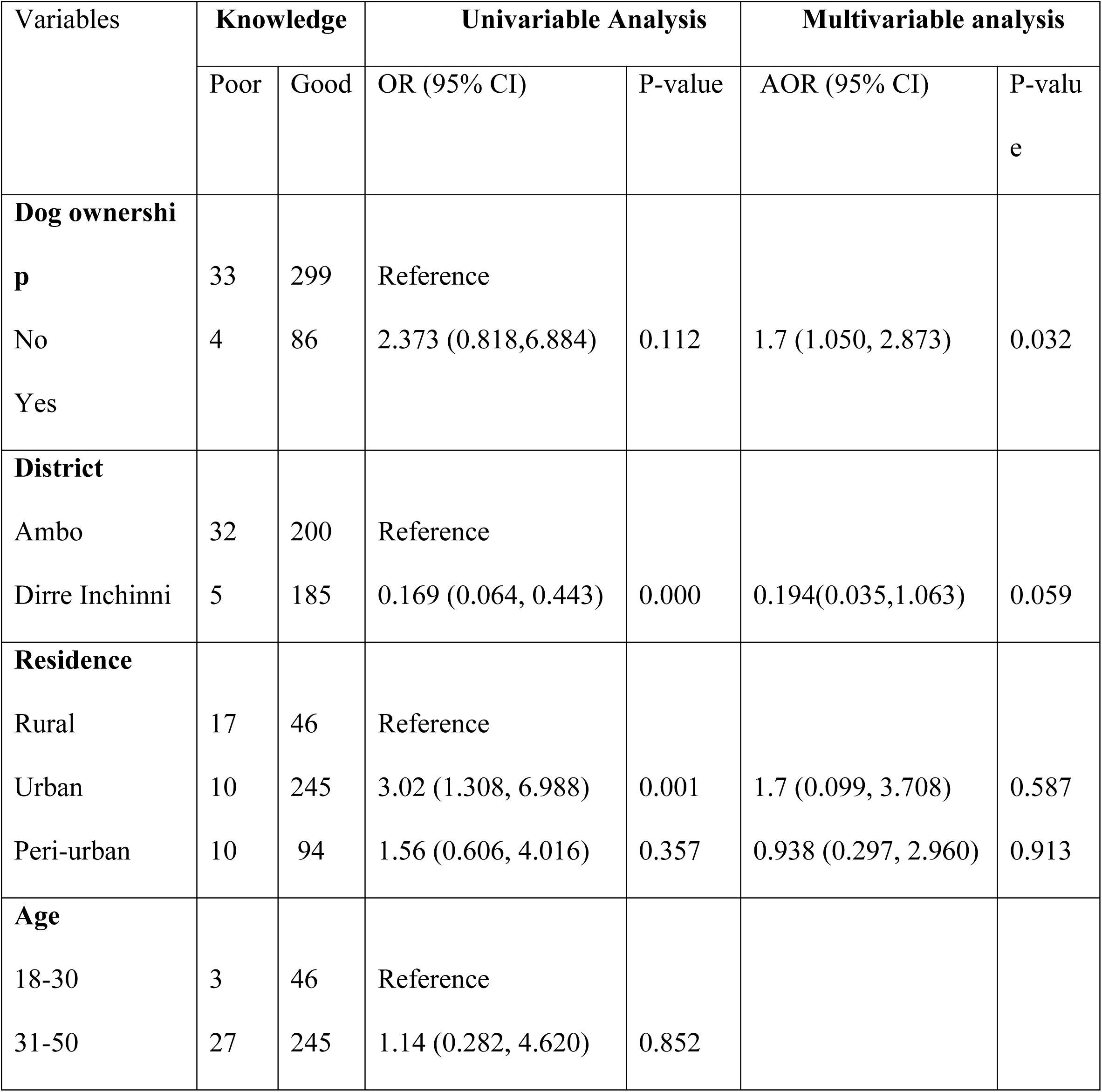
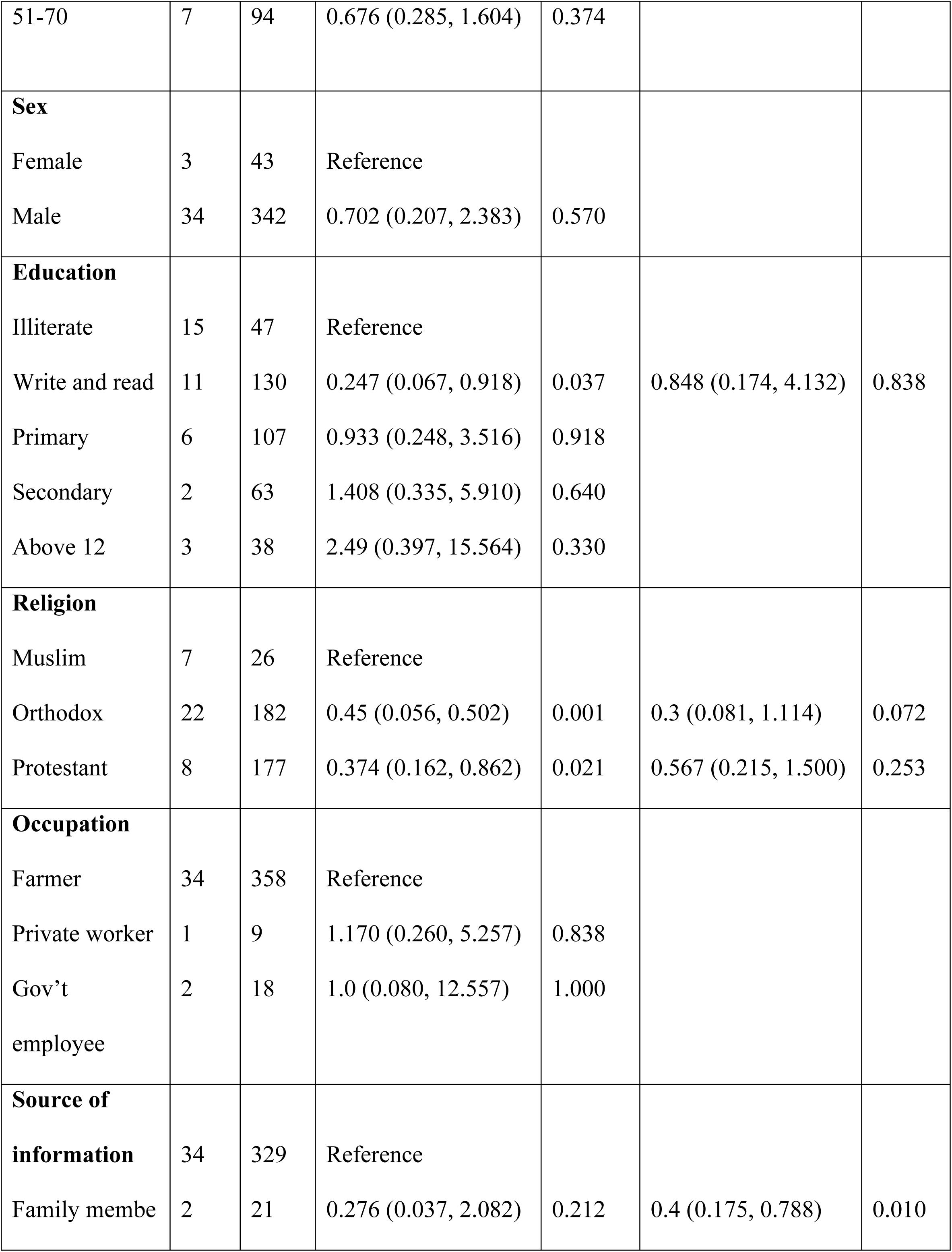
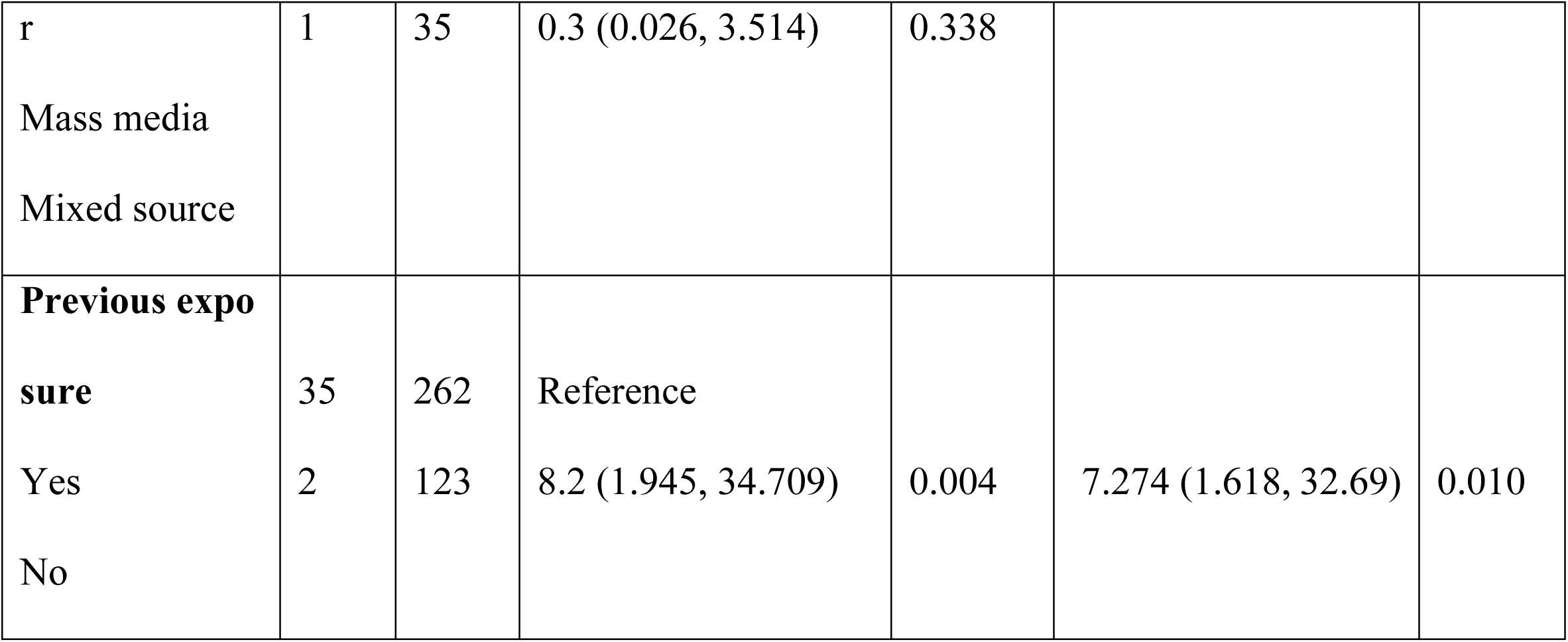
Univariable and multivariable logistic regression analysis between knowledge score on rabies and risk factors.

The results indicated that the majority of participants, 376 (89.1%), identified rabies as a health risk, and only 302 (71.6%) knew that vaccination could achieve rabies prevention. More than seventy percent of the study participants believed in the curative nature of the traditional healers. The majority, 276 (65.4%) of them, preferred traditional medicines over modern ones, as shown in (**Table 4)**.

**Table 4.** Community’s attitude indicating variables on rabies.

| Variables | Category | Number | Percent (%) |
| --- | --- | --- | --- |
| Rabies is a health risk | Yes | 376 | 89.1 |
|  | No | 46 | 10.1 |
| An exposed individual seeks medical evaluation immediately | Yes | 339 | 80.3 |
|  | No | 83 | 19.7 |
| Is rabies preventable by vaccination? | Yes | 302 | 71.6 |
|  | No | 120 | 28.4 |
| The frequency of rabies vaccination? | Annually | 69 | 16.4 |
|  | Don't know | 285 | 67.5 |
|  | Twice per year | 68 | 16.1 |
| Traditional healers can cure rabies? | Yes | 302 | 71.6 |
|  | No | 120 | 28.4 |
| Preferable treatment after exposure to rabies? | Modern drugs | 146 | 34.6 |
|  | Traditional drugs | 276 | 65.4 |
| Could rabies affect both humans and animals (zoonotic)? | Yes | 256 | 60.7 |
|  | No | 166 | 39.3 |

About 313 (74.2%) informants had a positive attitude while 109 (25.8%) had a negative attitude on rabies. Univariable logistic regression analysis showed that there was a statistically significant association between attitude score and dog owners (OR=4.5, 95% CI: 2.090, 9.609) and Dirre Inchinni residents (OR=1.8, 95%: 1.167, 2.815) as illustrated below in (**Table 5)**.

**Table 5.**
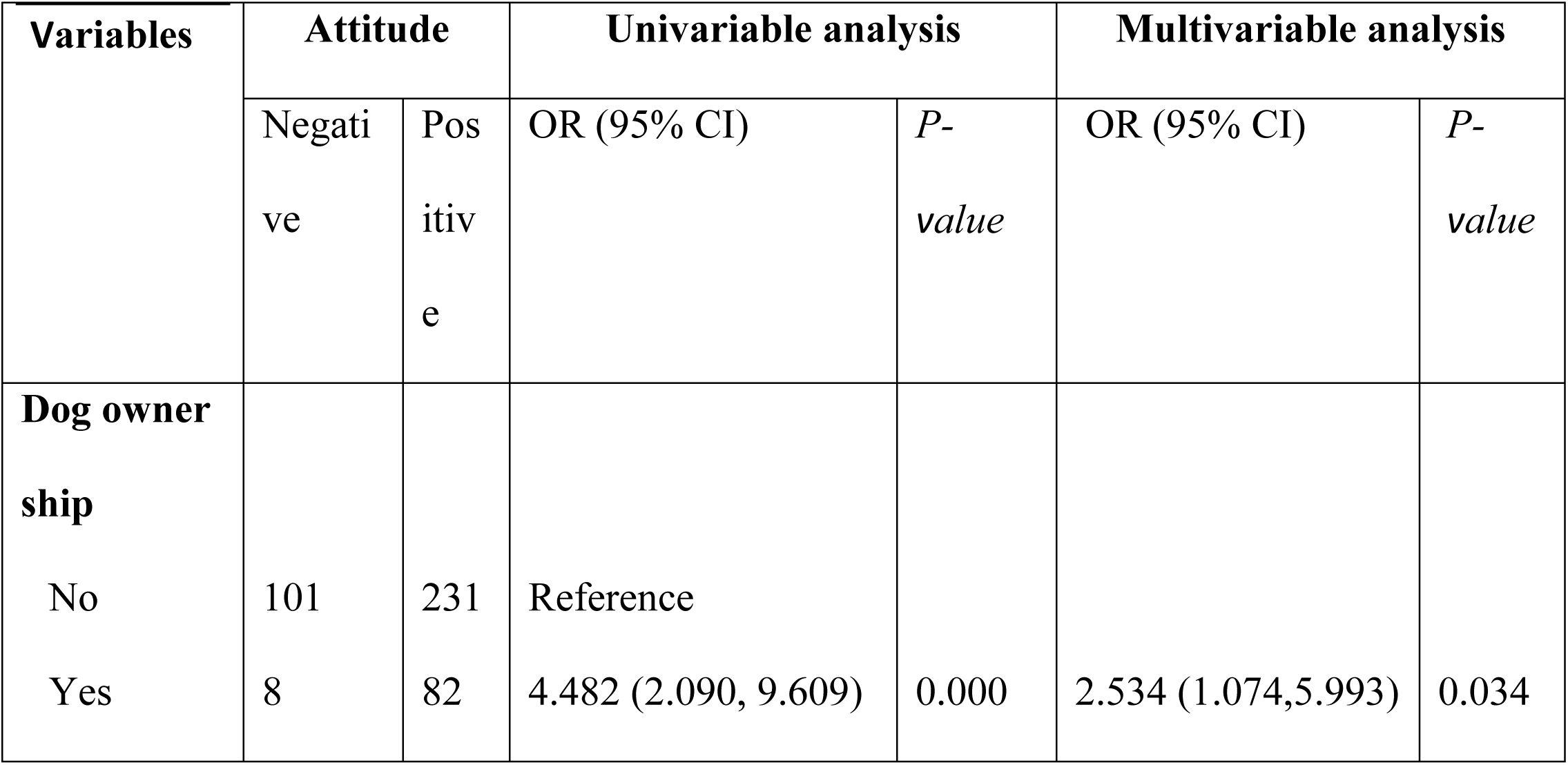
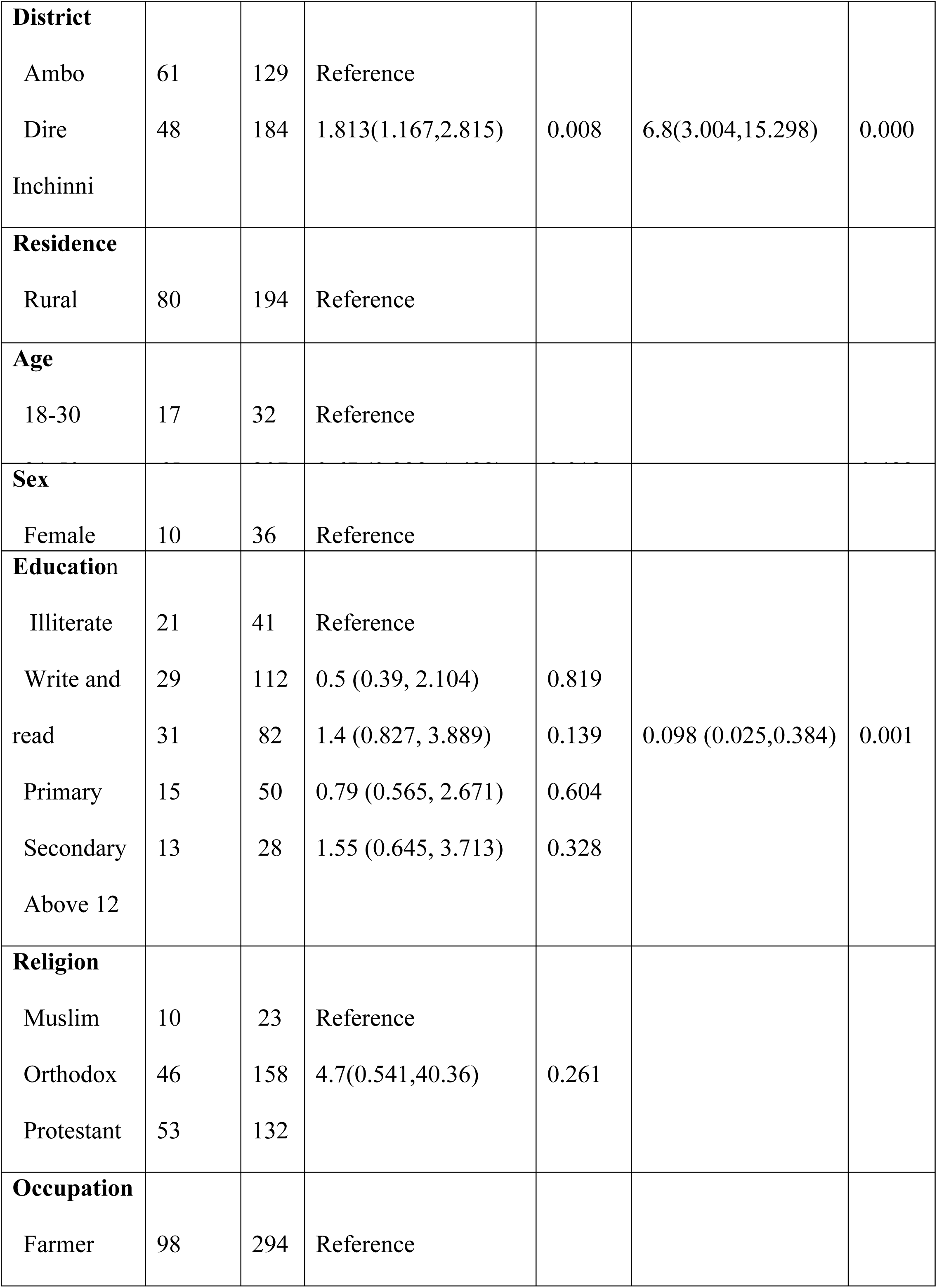
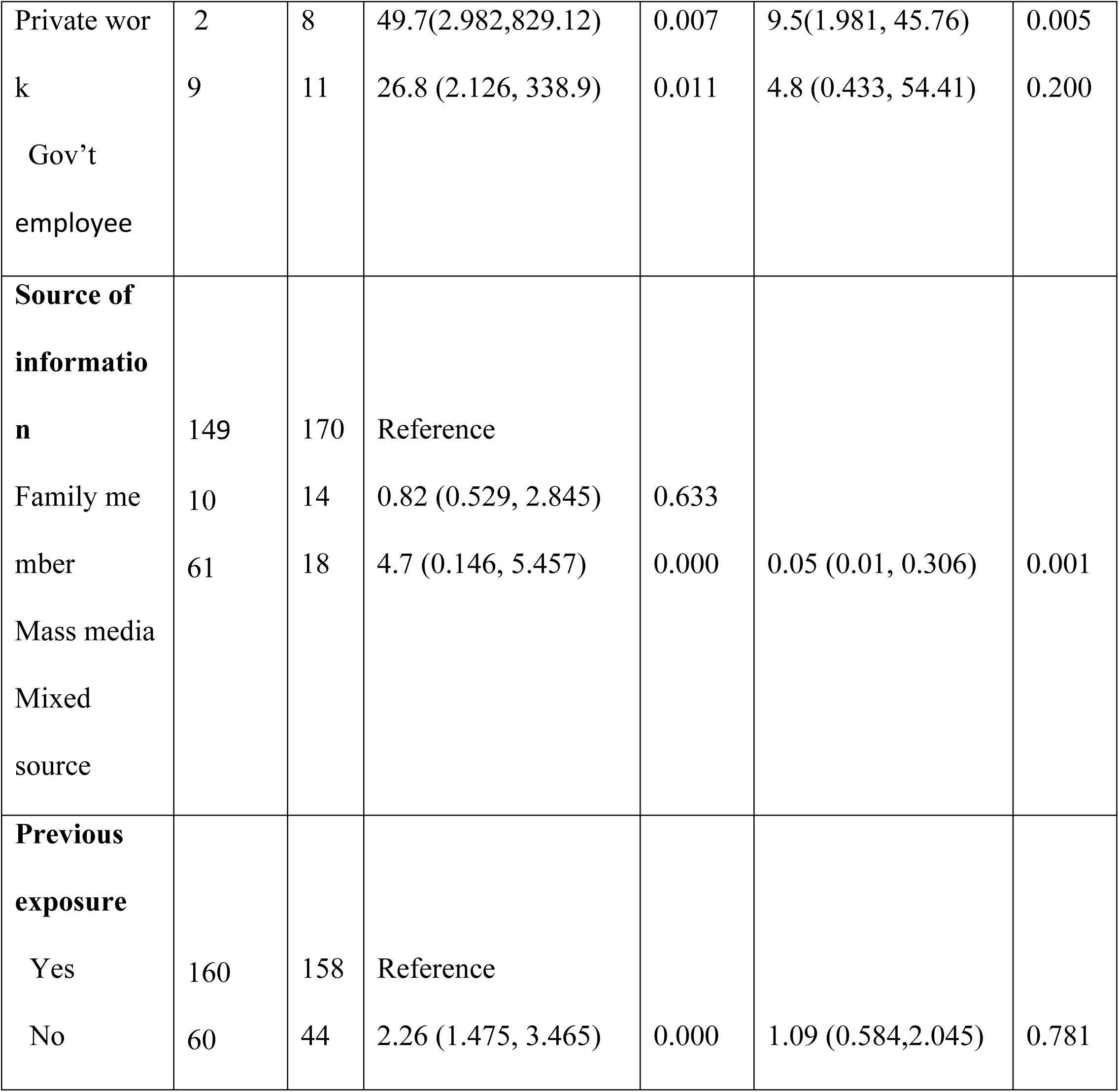
Univariable and multivariable logistic regression analysis between attitude score on rabies and risk factors.

The community’s practice indicator variables on rabies are displayed in (**Table 6**). The majority of 281 (66.6%) of the study participants, taking their diseased animal to the veterinary clinic was their preference in handling the case. More than half of the respondents, 259 (61.4%), considered that dead animals from rabies cases act as a source of infection for others.

**Table 6.**
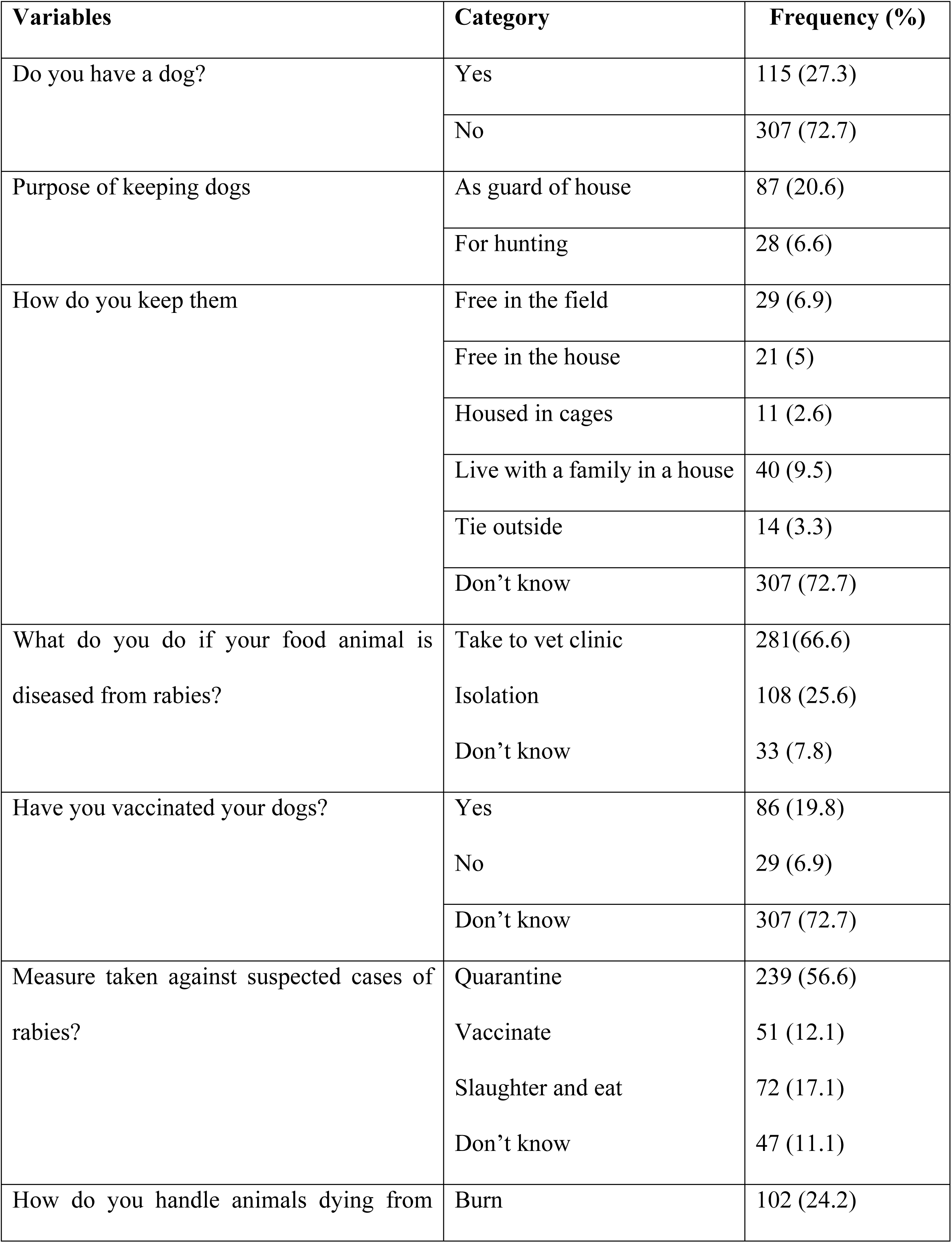
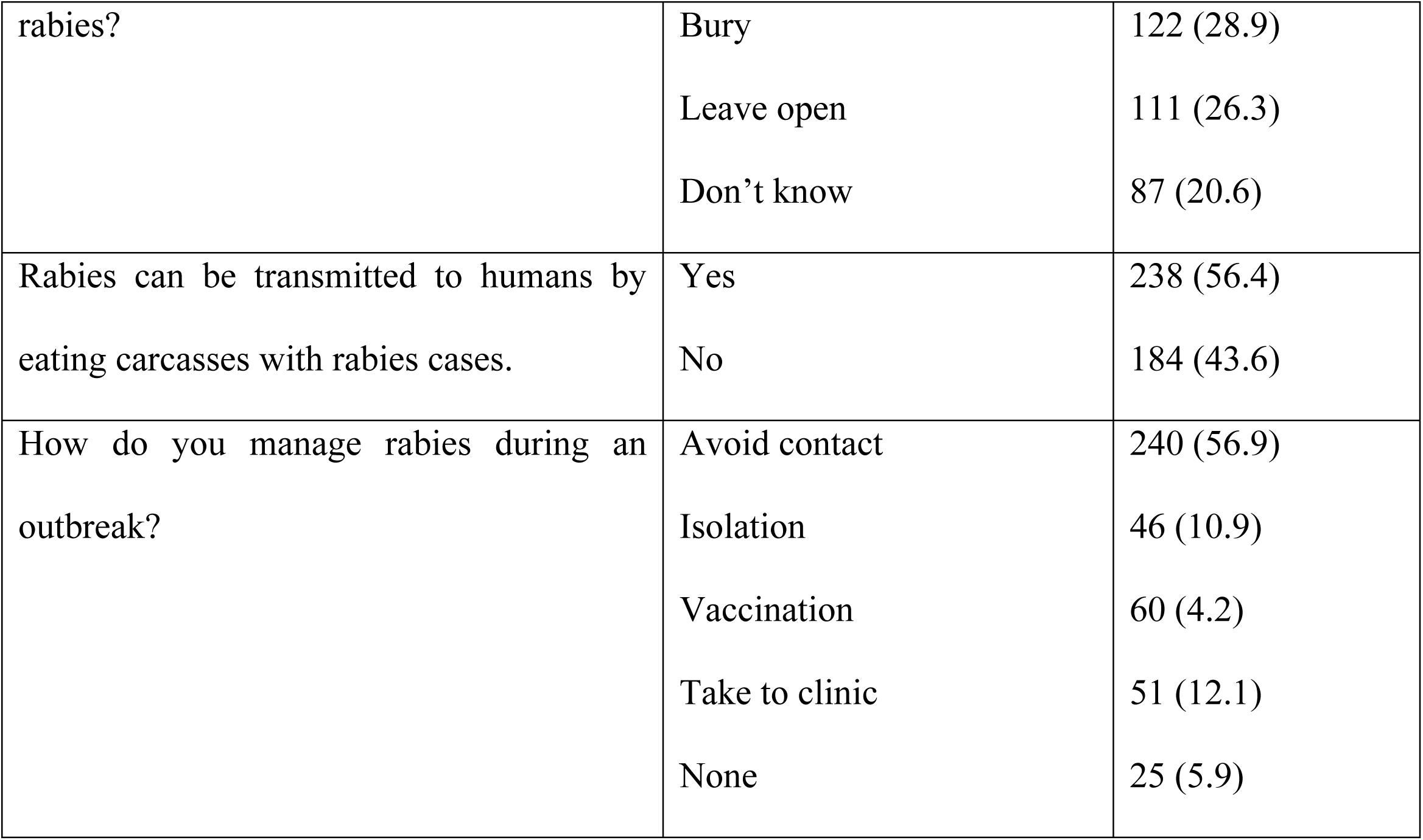
Community’s practice indicator variables on rabies.

In this study, about 342 (81%) respondents had good practice with rabies, whereas the remaining 80 (19%) had poor practice with rabies, as presented in (**Table 7**).

**Table 7.**
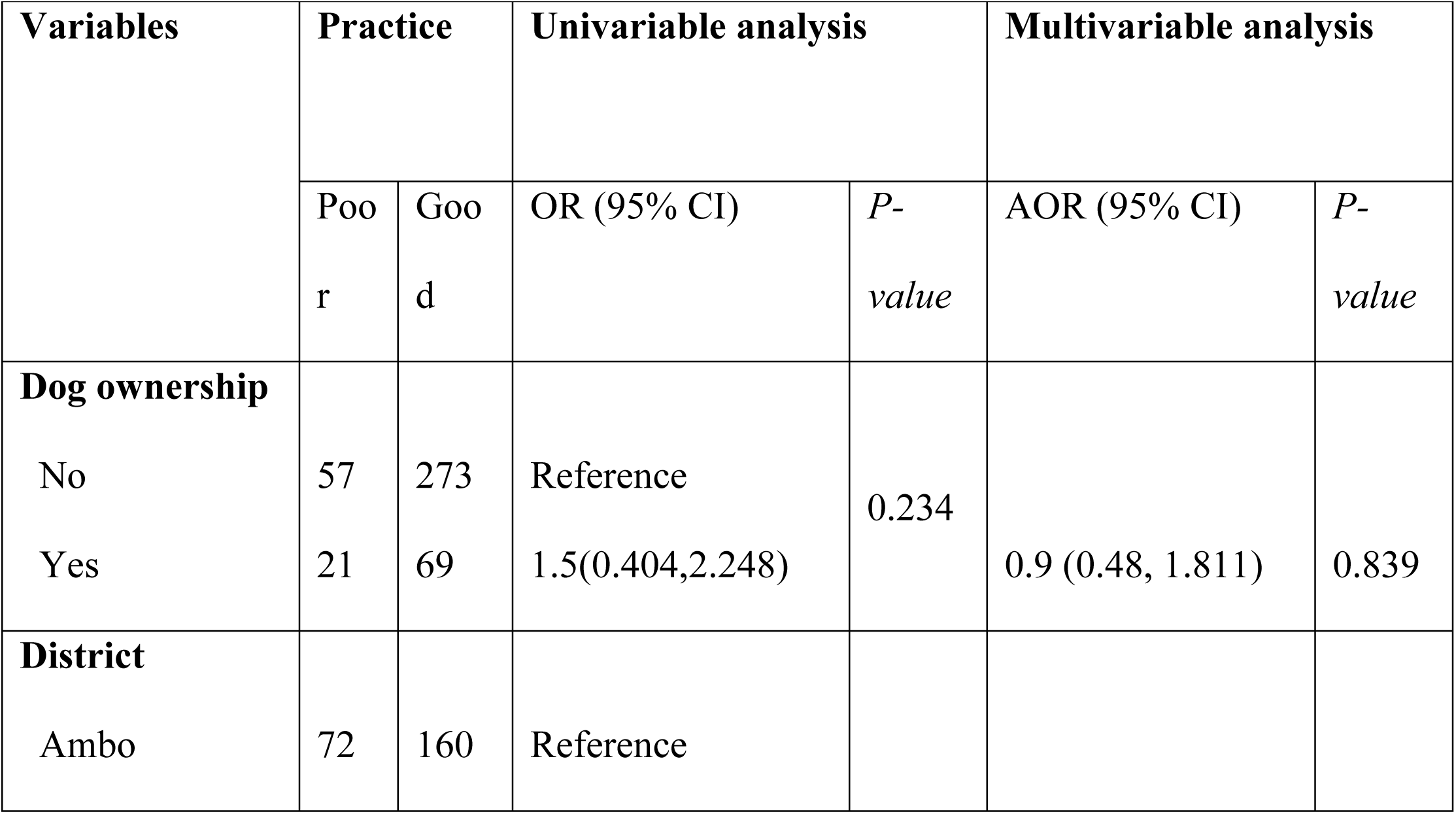
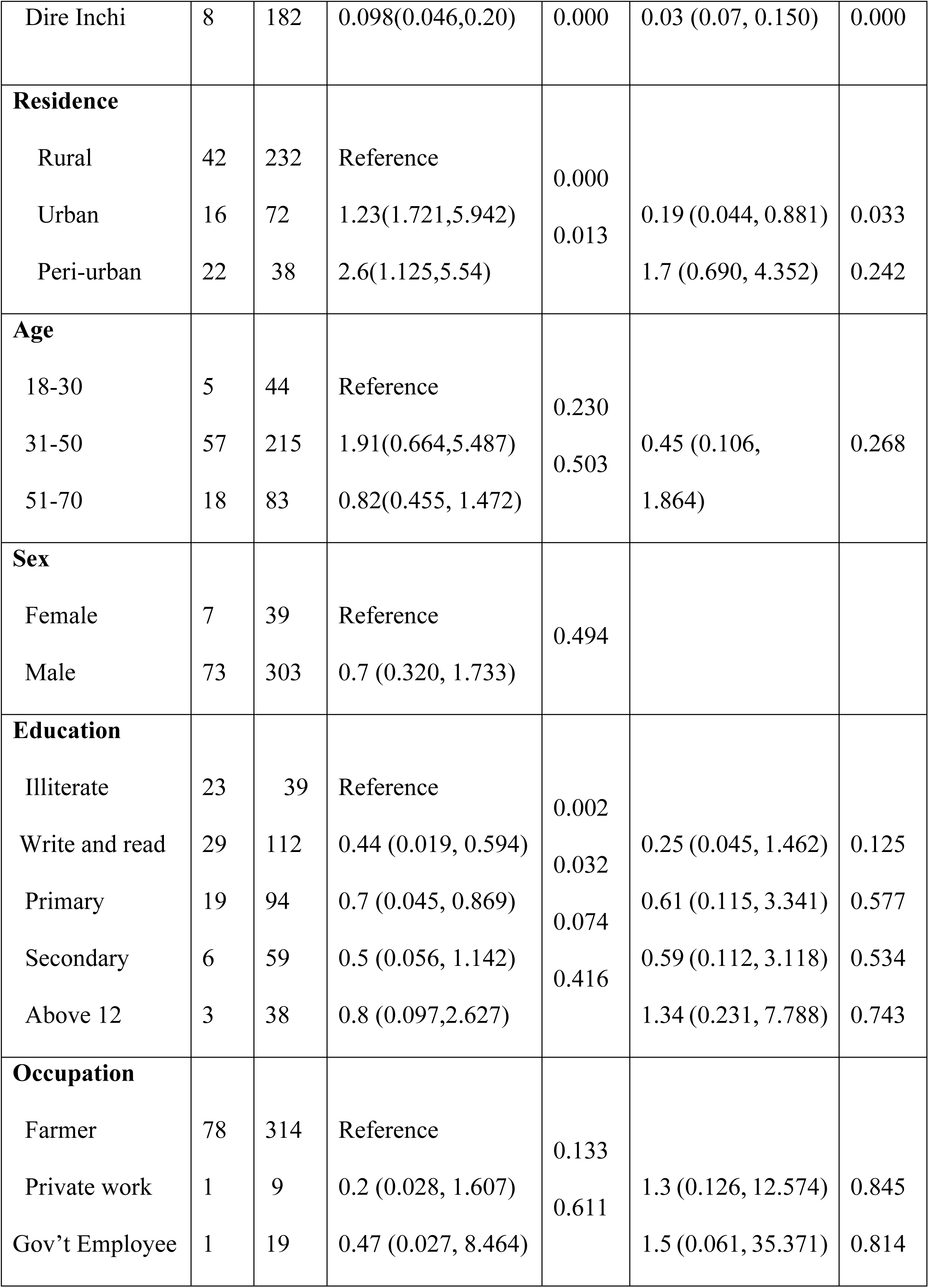
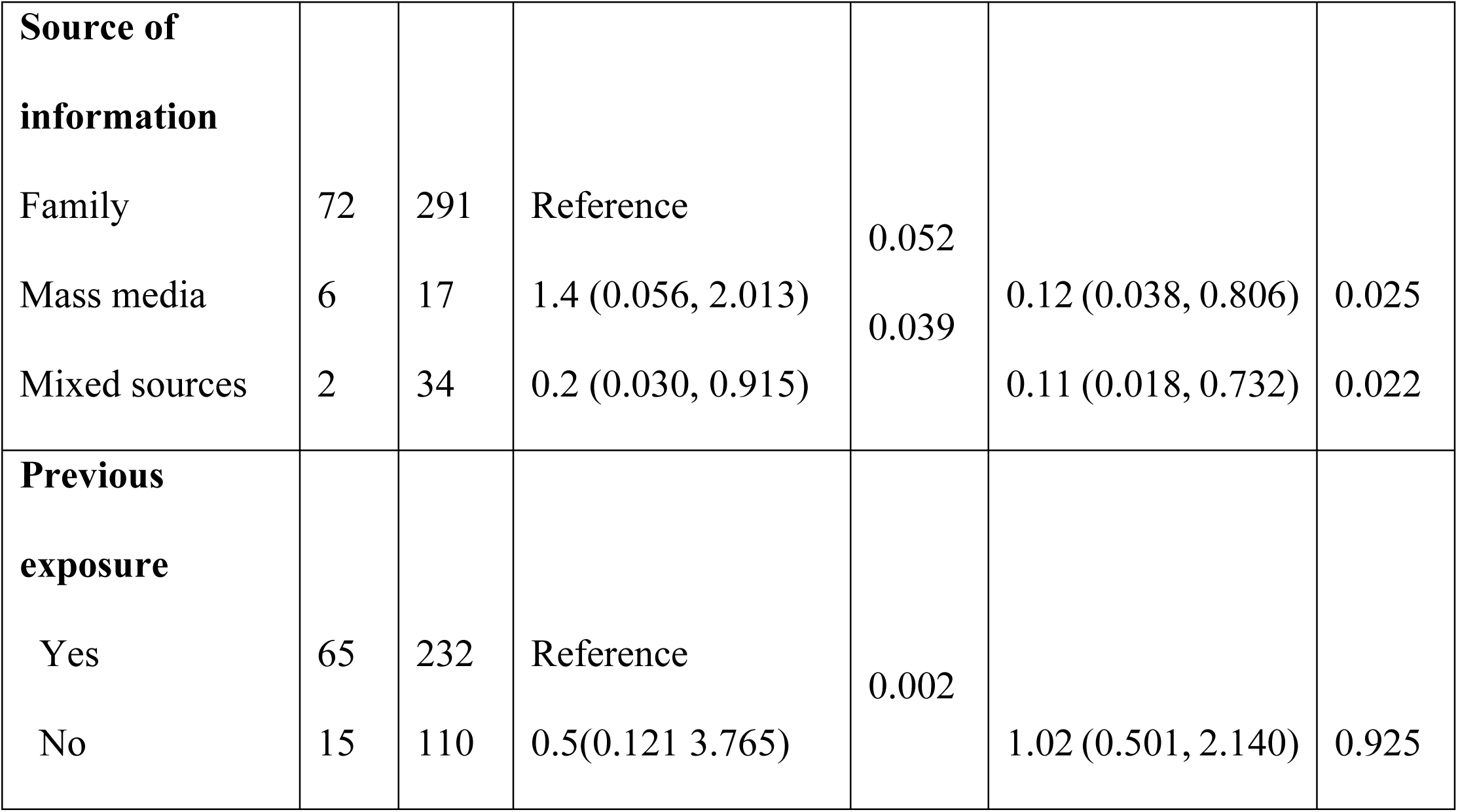
Univariable and multivariable logistic regression between practice score and risk factors on rabies in the study area.

### Retrospective cases of rabies in humans and animals

In this study, 579 rabies-suspected patients were recorded in the health centers of the districts from 2017 to 2021 in the hospitals’ casebooks. Rabies occurred with an average of 28.7 cases registered annually in the health centers of the districts. The majority of the cases were recorded in 2021, which was 139 (24%), and most of them were bitten by animals of unknown ownership, 431 (74.4%) on their legs 324 (60%). The retrospective data on rabies in the study districts is illustrated in (**Table 8)**.

**Table 8.** Incidence of suspected rabies cases in humans in the study area.

| **Variables** | **Number of cases** | **Percentage (%)** |
| --- | --- | --- |
| **Age** |  |  |
| Children (2-15) | 85 | 14.7 |
| Young (16-29) | 58 | 10 |
| Adult (30-43) | 190 | 32.8 |
| Elder (44-70) | 246 | 42.5 |
| **Sex** |  |  |
| Male | 438 | 75.6 |
| Female | 141 | 24.4 |
| **Residence** |  |  |
| Peri-urban | 170 | 29.4 |
| Rural | 204 | 35.2 |
| Urban | 205 | 35.4 |
| **Source of bite** |  |  |
| Canine (dog, cat) | 568 | 98.1 |
| Bovine (cattle) | 7 | 1.2 |
| Equine (horse, donkey, mule) | 4 | 0.7 |
| **Owner of the animal** |  |  |
| Known | 148 | 25.6 |
| Unknown | 431 | 74.4 |
| **Site of bite** |  |  |
| Body | 159 | 27.5 |
| Hand | 83 | 14.3 |
| Head | 7 | 1.2 |
| Legs | 324 | 56 |
| Multiple sites | 6 | 1 |

The number of rabies-suspected cases was highest in elders found above forty years, 246 (42.5%), followed by adults, 190 (32.8%) as shown in (**Table 8)**. The number of rabies-suspected individuals taking the post-exposure prophylaxis at district health centers varied accordingly. The maximum number of suspected rabies cases was recorded in 2021 (139 cases), and the lowest number was recorded in 2018 (72 cases).

The study indicated that the highest number of human rabies cases was documented in summer and autumn as compared to other seasons, as shown in (**Fig.4)**.

**Fig 4.**
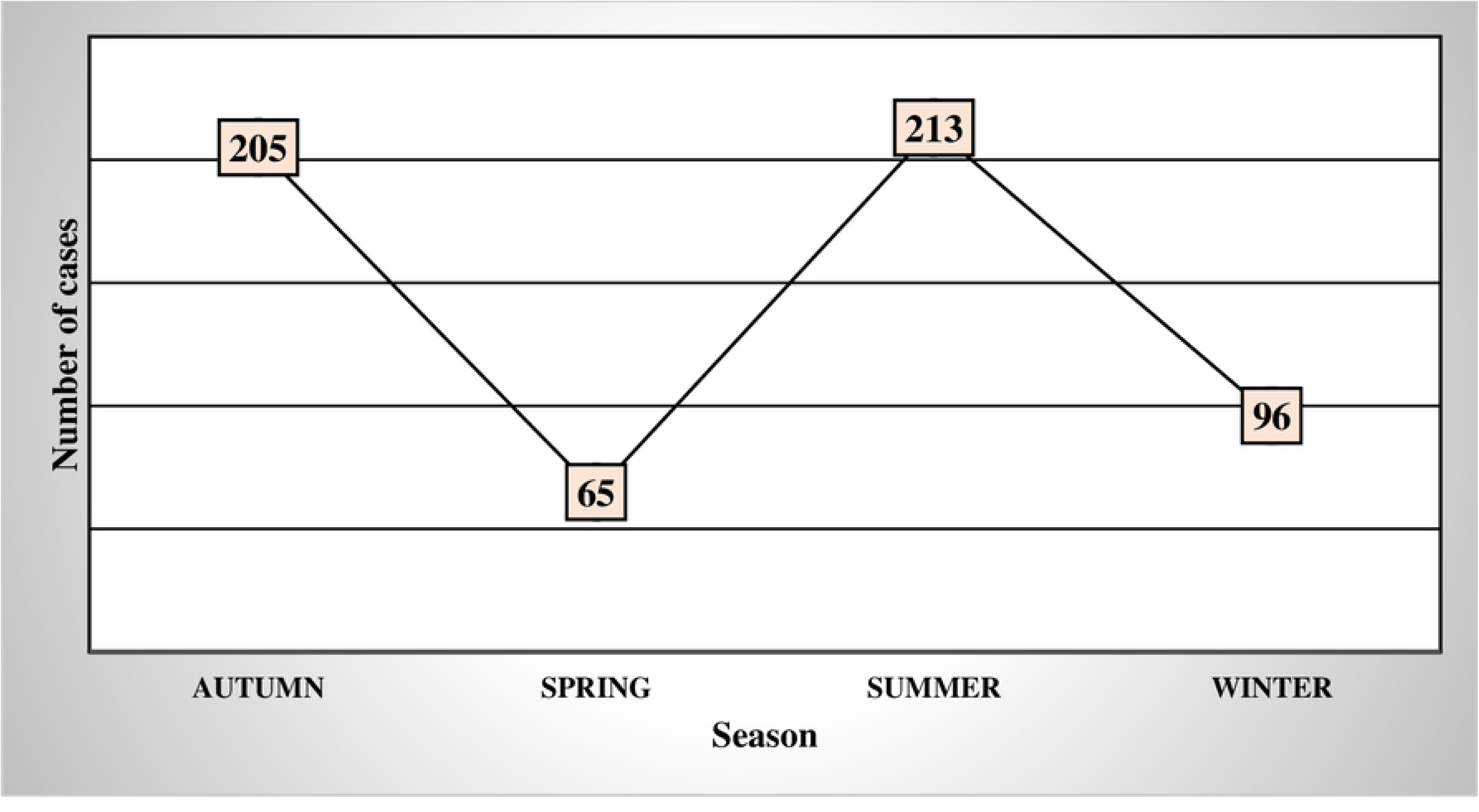
Seasonal distribution of rabies in humans

The occurrence of rabies was higher in humans than animals over the last five years in the study area. A total of 183 suspected rabies cases were registered in the case book at the district veterinary clinics during the study period. Of the total number of domestic animals, 92 (50.3%) cases were found in bovines, followed by canine and equine. The occurrence of rabies in the various district animal species is summarized in (**Table 9)**.

**Table 9.** Rabies cases in animals registered in the study area.

| Year | Total Cases |  | Suspected animal species for rabies |  |  |
| --- | --- | --- | --- | --- | --- |
|  | Cases | Percentage (%) | Bovine | Equine | Canine |
| 2017 | 26 | 14.2 | 15 | 6 | 5 |
| 2018 | 36 | 19.7 | 19 | 6 | 11 |
| 2019 | 53 | 29 | 25 | 10 | 18 |
| 2020 | 52 | 28.4 | 27 | 10 | 15 |
| 2021 | 16 | 8.7 | 6 | 3 | 7 |
| **Total** | **183** | **100** | **92** | **35** | **56** |

### Incidence of rabies in humans and animals in the study area

The incidence proportion of rabies in humans was higher in 2021 (31.1 cases per 100,000 population) and the lower incidence was recorded in 2018 (17.2 cases per 100,000 population) from retrospective data of the study periods (2017 to 2021) in the Ambo district. The incidence proportion of rabies in humans obtained from district health centers for five consecutive years (2017 to 2021) was higher in the Dire Inchinni district (32.7 cases per 100,000 population) than in the Ambo district (24.7 cases per 100,000 population) depicted as in (**Table 10)**.

**Table 10.**
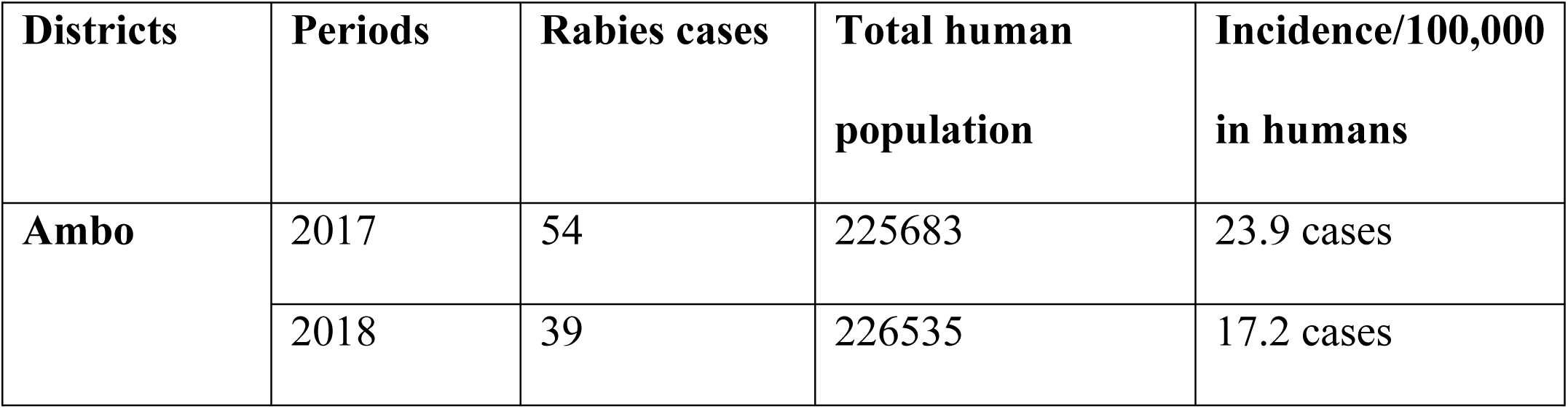
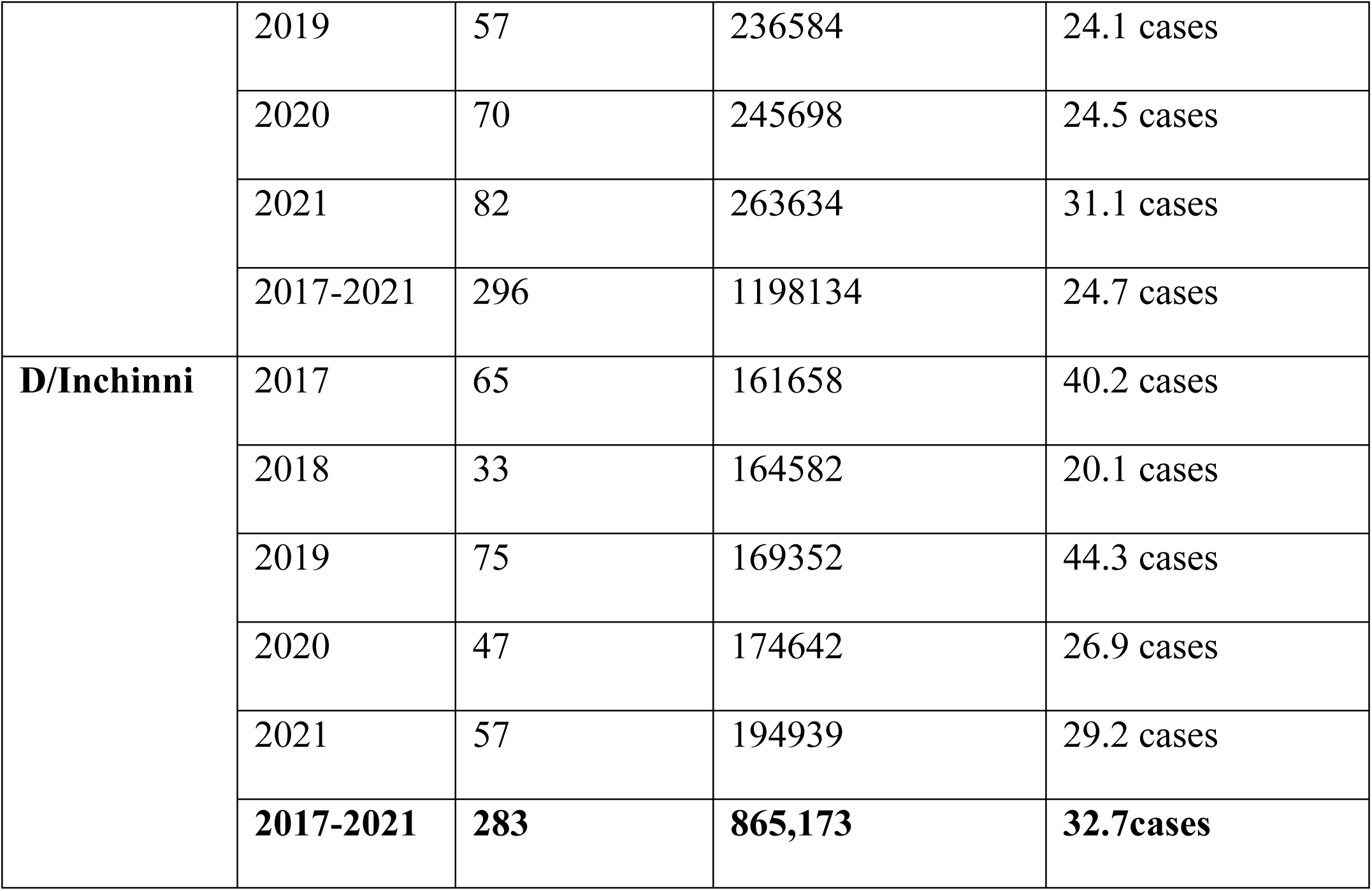
Incidence of rabies in humans in Ambo and Dire Inchini districts.

The incidence of rabies in animals was higher in 2020 (47.6 cases/100,000 populations) and lower in 2021 (10.9 cases/100,000 populations as indicated in (**Table 11**) below.

**Table 11.**
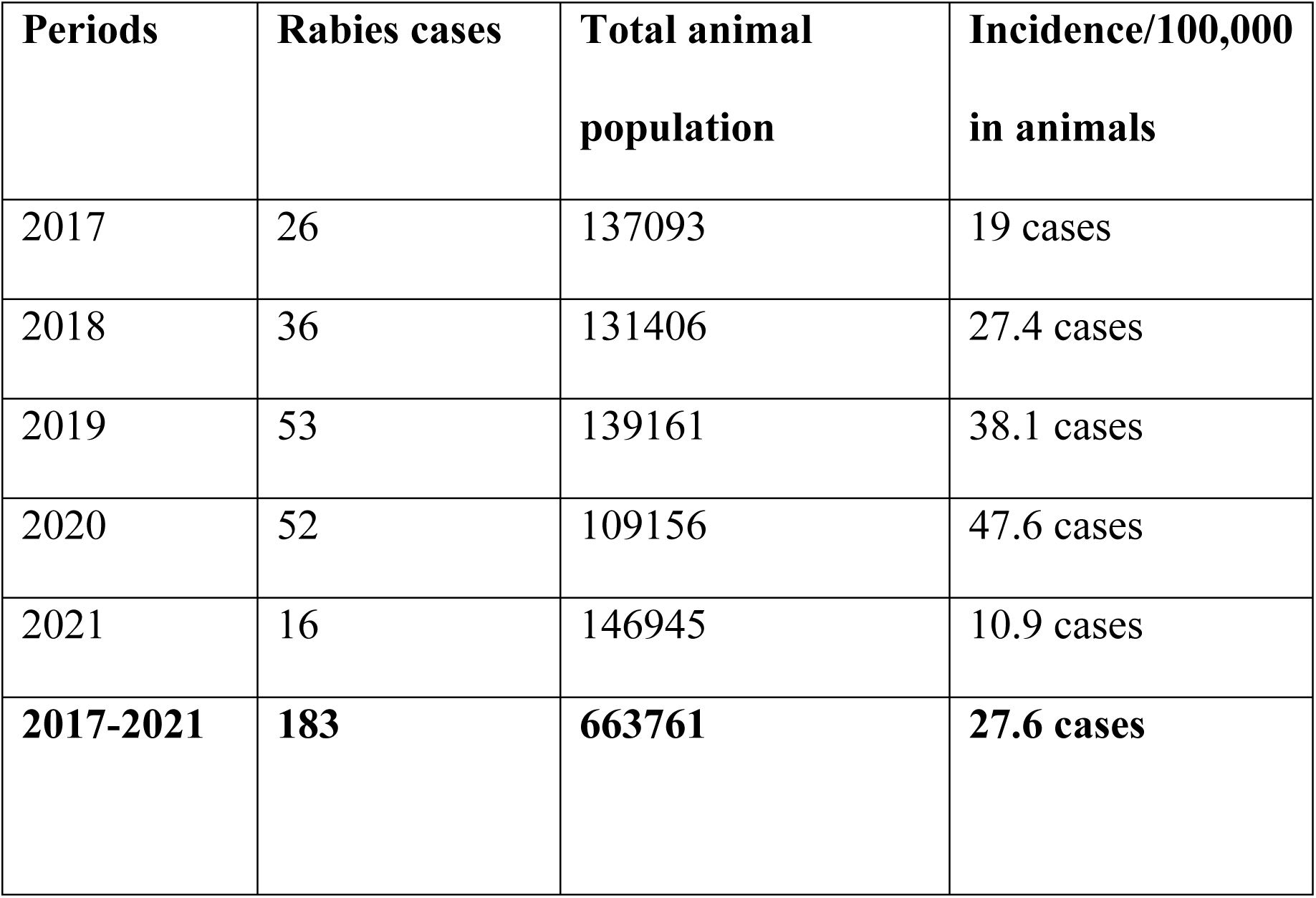
Incidence of rabies in animals in Ambo and Dire Inchini districts.

### Direct and indirect economic impact analysis of rabies in the study area

The total doses of PEP administered in the study area through the entire study period (2017-2021) are presented in (**Table 12**).

**Table 12.**
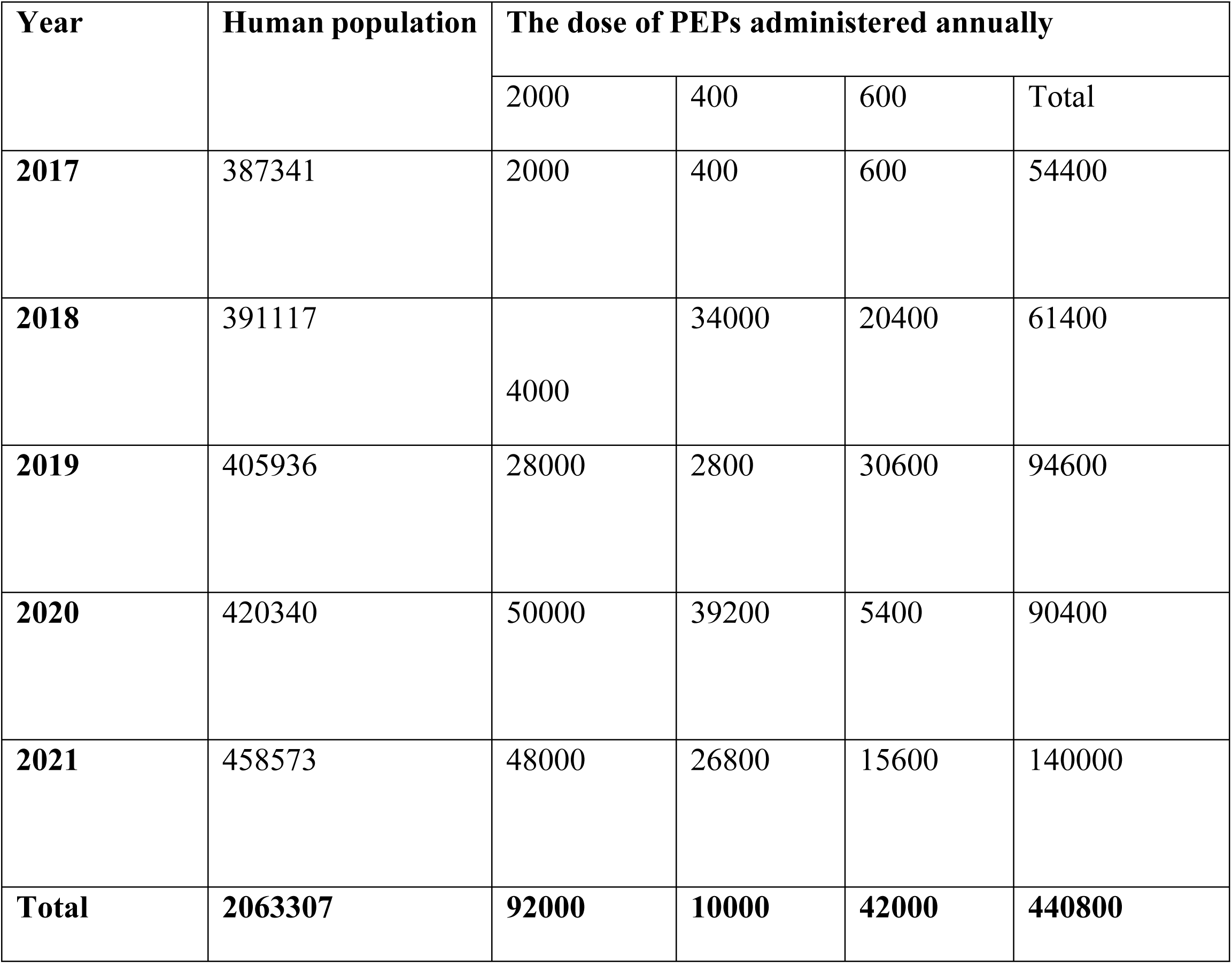
Human population and the total doses of PEPs administered in the study districts for 2017-2021.

The costs of rabies were classified into two direct cost categories those are healthcare costs reflected by the expenditures for rabies vaccine, antibiotics, tetanus immunizations, and disinfection, and non-healthcare costs as the expenses for transport, food, and accommodation while seeking PET Rabies has posed significant economic impacts in the study districts—the total costs associated with human rabies in the area are depicted in (**Table 13**).

**Table 13.**
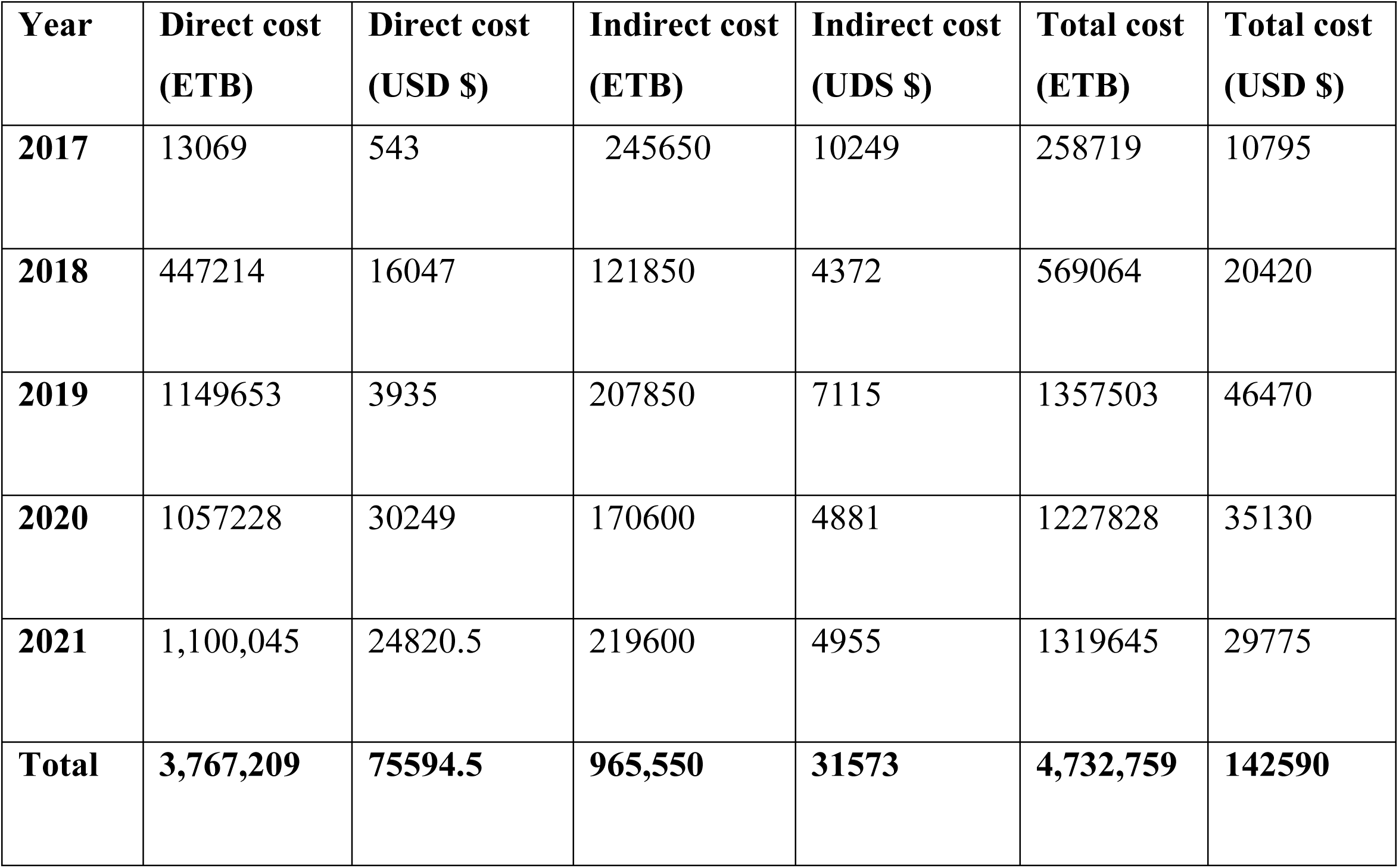
Total cost (USD) associated with human rabies in the study area from 2017-2021.

## Discussion

As a developing nation, Ethiopia has a high rabies endemicity [14]. In the current study, 395 (93.6%) respondents were heard about rabies from different sources. The finding is relatively aligned with the finding of the survey done in Arsi town, Ethiopia, from different community members by [9] who reported that 97.9% of participants knew about rabies. However, it was higher than the reports of [19] in Tanzania, [20] in Guatemala, and [21] in Western India, who reported a knowledge score of 49%, 27%, and 0% respectively. The difference could be associated with the awareness level of the community, educational status, and information access. In the study, about 338 (80.1%) informants obtained information from informal sources. Similarly, the Gondar and Mekele study indicated that most of the respondents acquire information about rabies from informal sources [8,22], respectively. The result did not agree with the findings of [23], in which half of the respondents (48%) were informed about rabies from several sources other than family and friends. This might be due to the frequency of contact among the community and the culture of discussing issues in their locality.

From all study participants, only 115 (27.3%) individuals owned a dog as a domestic animal in their house. Nevertheless, there was good awareness of variables related to rabies infection, the zoonotic and fatal nature of rabies, and prevention methods through vaccination. The result aligns with the finding of [24], who reported that around 690 (76.8%) of participants did not own dogs but had a good awareness of rabies. These high levels of awareness are due to this population’s greater access to several means of communication and education (radio, television, mobile phones, and educational establishments).

Dog vaccination practice was generally deficient and nonexistent in both districts of the study area. Even if the community awareness was better, lack of access, lack of vaccines, and absence of veterinary clinics in the area were raised as problems by the respondents. Among the study participants, 29 (6.9%) of household heads had vaccinated their dogs, which is higher than the study conducted in Jimma by [25], which was 4.8% and lower than the study done in Gondar by [26] reported 42%. This could be attributed to several factors, including the availability of vaccines, the study time, and information-sharing habits in this study area among the community.

Household heads with good knowledge about rabies were 91.2%, which was higher than studies conducted in Addis Ababa, Bahir-Dar, Gondar, and Debretabor, who reported 83%, 84.9%, 60.1%, and 90.8%, respectively [11,12,24,26]. The difference could be due to low health promotion, particularly regarding rabies in this study area. Among the household heads in this study, 74.2% had a positive attitude about rabies, which was lower than a study conducted in Indonesia by [27) reported 96% and [24] who forwarded 98.6% of desirable attitudes. The lack of more information about rabies might explain the difference in the study site. Moreover, the attitude score of the current finding is greater than the study conducted in Debark by [28] reported 40.6%. This may be due to differences in information levels among the community, which could bring a difference in the awareness of study participants.

In the present study, 392 (92.9%) participants knew about the transmission of rabies from animal to human. This finding was higher than the result of [11], who reported 21.4% from Bahir Dar town. The possible reason for this could be due to the availability of different host ranges, the level of awareness, and the educational status of the community. Most 386 (97.7%) respondents considered rabies to be a fatal disease. The result was higher than [9], who reported 21.9% in Mozambique. However, this finding was discordant with the study conducted in different areas of Ethiopia ([11,29]. This could be associated with their previous exposure to the disease and the awareness level of the community.

The results also showed that most communities in the study area were familiar with general information on rabies through 91.2%, 74.2%, and 81% knowledge, attitude, and practice scores, respectively. This aligns with the study on rabies conducted in different parts of Ethiopia [14,30]. This may be due to rabies being considered an endemic and well-known disease in Ethiopia for extended periods. In this study, 60.7% of respondents knew that rabies could affect humans and other domestic animals, which is in line with [29]. However, [11] reported a lower result (21.4%) from Bahir Dar, and a higher impact (71.9%) was also reported in the city of New York by [31]. The possible reasons for this could be the availability of different host ranges, the level of awareness, and the educational status of the community.

Respondents’ knowledge scores on rabies were significantly associated with dog ownership (OR=1.7, 95% CI: 1.050, 2.873), and previous exposure to rabies (OR= 7.3, 95%CI: 1.618, 32.69). The statistically significant difference in knowledge score between dog owners, prior exposure, and non-dog owners could be attributed to the fact that dog owners have had a better chance to know more about dog and dog disease. Many scholars supported this finding and mentioned awareness level is an essential tool for controlling rabies [14,22]. Regarding attitude, dog owners are 2.5 times more likely to have a positive attitude towards rabies than those who don’t own dogs. The result is in line with the findings of [22], who found that private workers have better positive attitudes.

This study revealed that 579 total human rabies-suspected cases were registered in the health facilities, including hospitals and health centers, from 2017 to 2021 in the study districts. This finding is significantly lower than that of a study conducted in northwestern Amhara [32], Northwestern Tigray by [33], and Jimma Zone [25], where 924, 2180, and 2302 human rabies exposure cases by dog bite were reported, respectively. This might be due to increased awareness of suspected human rabies by dog bites and improvement in health-seeking behavior, which might have increased in the report of suspected human rabies cases by a dog bite.

In the present study, the number of rabies cases from the retrospective data was highest in 2021 compared to other study periods. Most rabies-suspected individuals 388 (92%) took nerve tissue vaccine (NTV) as post-exposure prophylaxis. The finding is in line with the study done by [12] in Addis Ababa and surrounding towns, in which most of the patients took nerve tissue vaccines. This could be due to the vaccine’s cost and availability in Ethiopia’s health centers.

More rabies-suspected cases were reported in humans (579 cases) than in animals (183 cases) over the last five years of the retrospective survey. This might be associated with the weak disease reporting system in the veterinary sector. Compared to the present study, more human rabies cases (2798 cases) were reported in the Tigray region [34]. A survey by Mengistu et al.[35] and his colleagues indicated human rabies cases (4729) and 44 deaths in the Tigray region from 2009 to 2012. This could be due to the high dog population, the poor trend of the communities to vaccinate their dogs, and the lack of awareness about the disease.

Most rabies-suspected cases, 568 (98.1%) were due to bites of rabid dogs of unknown ownership. This is consistent with the study done in Jimma Town by [25], which indicated that a significant proportion of the interviewed households (97.2%) suggested rabies is transmitted to humans when they are bitten, scratched, or licked by rabid dogs. Another study by [36] also reported that 97% of humans in Kenya used post-exposure treatments to rabies due to dog bites.

The majority 437 (75.5%) of rabies-suspected individuals were male victims. This aligns with the study in Addis Ababa by [12], in which most exposed individuals were male. This study showed that most individuals get bitten on their legs 324 (56%) and hands 83 (14.3%). The finding aligns with the study done at Ambo town by [16] and contradicts the survey done by [12], who reported that most of them were bitten on their hands rather than legs. This could be due to infants and schoolchildren who play closely with pets at home and even on the streets being more likely to be exposed than adults.

In the case of economic analysis, over 3,767,209 ETB (75594.5$) were lost due to rabies in the study area as a direct cost, and around 965,550 ETB (31,575$) were lost as an indirect cost in rabies patients in this study period. Finally, the total cost expensed on rabies by all individuals who had taken rabies prophylaxis was 4,732,759 ETB (142,590$). The incidence of rabies in humans obtained from district health centers for five consecutive years (2017 to 2021) was higher in the Dire Inchinni district (145.2 cases per 100,000 population) than in the Ambo district (112.3 cases per 100,000 population). The result of the current study agrees with the findings of [35]. Those who reported the highest number of rabies cases in selected districts of the Tigray Region disagree with the finding of [37], who reported 0.05 cases per 100,000 population; this could be due to differences in human population and time of study.

## Study Limitations

Because of the limited number of districts in the study, the findings might not represent and reflect the accurate geographical distribution and incidence of rabies. It is difficult to determine accurate incidence of rabies from retrospective data because of underreporting, misdiagnosis, incomplete records, differences in the surveillance system, and focus on human cases and overlooked animal cases. Also, it could be associated with the affected animals are in a remote area, and there is a lack of knowledge or access to veterinary care. The actual number of cases is therefore probably underestimated. Poor recording practices and delayed outbreak investigation approach might result in incomplete or inconsistent data in animal and human rabies cases. The other limitation of this study was the variability among the district’s health facilities (hospitals, health centers, and veterinary clinics) in the study area. Despite the study’s limitations, the study is still significant in showing the level of knowledge, attitude, and practice in the study area. The finding elicits further awareness creation campaigns through different training and education for effective prevention and control of rabies in the country.

## Conclusion

According to the current study, rabies is a well-known and endemic disease in the study area, in both humans and animals. The knowledge score on rabies was highly associated with dog ownership, source of information, and previous exposure. Among the risk factors associated with an attitude score are dog owners, private workers, and education level. There was a lack of consistent understanding of the disease among the participants. The study also showed that the participants did not receive regular, sufficient, or ongoing health education regarding the disease. Even though the disease threatens farmers’ economies through the loss of animals, no action has been taken to combat its spread and transmission. Therefore, based on the above conclusion, the following recommendations were forwarded. Greater engagement of the veterinary and health sectors is needed to ensure the availability of preventative services, awareness programs, facilitation of vaccines, and regular vaccination of rabies in the study area. The case recording system of both human health centers and veterinary clinics should be upgraded to a computer-based database system to obtain accurate and comprehensive data when required. The government should develop effective, well-organized control measures for identified risk factors.

## Data Availability

Data cannot be shared openly but is available with reasonable requests from authors.

## Acknowledgments

We would like to thank the administrative bodies and veterinary experts of Ambo and Dire Inchini District Agriculture Office, the participants involved in the interview, and the Development Agents (DAs) for their cooperation during data collection from the study sites.

